# Seasonal Dynamics of *Anopheles stephensi* and its Implications for Mosquito Detection and Emergent Malaria Control in the Horn of Africa

**DOI:** 10.1101/2022.09.28.22280458

**Authors:** Charles Whittaker, Arran Hamlet, Ellie Sherrard-Smith, Peter Winskill, Gina Cuomo-Dannenburg, Patrick G.T. Walker, Marianne Sinka, Samuel Pironon, Ashwani Kumar, Azra Ghani, Samir Bhatt, Thomas S. Churcher

**Affiliations:** MRC Centre for Global Infectious Disease Analysis & Abdul Latif Jameel Institute for Disease and Emergency Analytics, School of Public Health, Imperial College London, London, UK; Department of Biology, University of Oxford, Oxford, UK; Royal Botanic Gardens Kew, Richmond, Surrey, UK; United Nations Environment Program World Conservation Monitoring Centre, Cambridge, UK; Vector Control Research Centre, Indira Nagar, Puducherry, India; Section of Epidemiology, Department of Public Health, University of Copenhagen, Copenhagen, Denmark

**Keywords:** *Anopheles stephensi*, malaria ecology, urban malaria, population dynamics, seasonality, epidemiology

## Abstract

Invasion of the malaria vector *Anopheles stephensi* across the Horn of Africa threatens control efforts across the continent, particularly in urban settings where the vector is able to proliferate. Malaria transmission across Africa is primarily determined by the abundance of dominant vectors, which often varies seasonally with rainfall. However, it remains unclear how *An.stephensi* abundance changes throughout the year, despite this being a crucial input to surveillance and control activities. We collate longitudinal catch-data from across its endemic range to better understand the vector’s seasonal dynamics and explore the implications of this seasonality for malaria surveillance and control across the Horn of Africa. Our analyses reveal pronounced variation in seasonal dynamics, the timing and nature of which are poorly predicted by rainfall patterns. Instead, they are associated with temperature and patterns of land-use, with seasonality frequently differing between rural and urban settings. Our results show that timing entomological surveys to coincide with rainy periods is unlikely to improve the likelihood of detecting *An.stephensi*. Integrating these results into a model of malaria transmission, we show that timing indoor residual spraying campaigns to coincide with peak rainfall offers little improvement in reducing disease burden compared to starting in a random month. Our results suggest that unlike other major malaria vectors in Africa, rainfall may be a poor guide to predicting the timing of peaks in *An.stephensi*-driven malaria transmission. This highlights the urgent need for longitudinal entomological monitoring of the vector in its new environments given recent invasion and potential spread across the continent.

## Introduction

There has been an estimated 40% reduction in the burden of malaria since 2000, predominantly due to significant scale-up of control interventions^1^. Increasing urbanisation of Africa’s human population (31% to 43% between 1990 and 2018, with >60% expected to live in urban areas by 2050^2^) is also thought to have indirectly contributed to reductions in disease burden. Previous work has found significantly lower Entomological Inoculation Rates (EIR) in urban compared to rural settings^3,4^. This is thought to be underpinned by factors including differences in housing quality^5,6^, reduced suitability of habitats for *Anopheles* breeding in urban settings^7–9^, better access to treatment^10^, and higher population densities leading to lower mosquito-to-human ratios (and reduced transmission)^11^. Whilst these trends are not always consistently identified^12,13^ (including previous work showing some vectors can adapt to urban environments^14^), increasing urbanicity across Africa is anticipated to complement planned scale-up of malaria control interventions aimed at achieving the targets outlined in the World Health Organization’s 2030 Global Technical Strategy for Malaria^15^.

This beneficial impact of increasing urbanization on malaria burden is contingent on urban settings remaining areas of comparatively low transmission. This is currently under threat in Africa because of the invasion and establishment of *An. stephensi*, a malaria vector that is potentially capable of thriving in urban areas of the continent^16^. There are three known forms of the species (“type”, “intermediate” and “mysorensis”) found across its native range in South Asia. The mysorensis form is predominantly found in rural settings, is highly zoophilic and typically possesses a low vectorial capacity^17^. By contrast, the type and intermediate forms represent efficient vectors capable of transmitting both *Plasmodium falciparum* and *Plasmodium vivax*^18–20^ in urban environments. This ability to proliferate in urban locations distinguishes this species from other malaria vectors in sub-Saharan Africa, and is thought to be underpinned by an increased tolerance for breeding in polluted water sources^21^, and superior ability to utilise the purpose-built water storage tanks present in many urban settings^22,23^.

The African invasion by *An. stephensi* was first reported from Djibouti City in 2012^24^ and has since been recorded in Ethiopia^18,25^, Sudan^26,27^, Somalia^28^ and Somaliland^29^, with recent work highlighting suitability of the continent’s largest population centres (where >100 million individuals live) as a habitat for this species^16^. Whilst causality has yet to be established, emergence of *An. stephensi* is thought to have contributed to resurgence of malaria transmission in Djibouti (10-fold increase in cases 2013-2019), highlighting the potential threat that this vector poses to malaria control across the Horn of Africa^30^ and the continent more generally^31^.

Despite the significant public-health this vector poses, substantial uncertainty remains in how its establishment might influence malaria dynamics in the region, particularly in the (predominantly urban) settings where the disease is currently largely absent. A key driver of this will be the vector’s seasonal dynamics. Mosquito populations may show marked variation in seasonal abundance, often exhibiting substantial annual fluctuations in size that drive the temporal profile of disease risk. The efficacy of many malaria control interventions (such as seasonal malaria chemoprevention^32^ (SMC), indoor-residual spraying^33^ (IRS) or larval source management^34^ (LSM)) depends on optimally timing their delivery relative to seasonal peaks in vector abundance. A better understanding of the seasonality of *An. stephensi* across its current range will help guide entomological monitoring and surveillance activities in areas of possible invasion and have material consequences for the effective control of *An. stephensi* driven malaria transmission.

Here we systematically collate longitudinal catch data for *An. stephensi* across its endemic range to better understand these dynamics. Our results highlight pronounced variation in the extent and timing of seasonality (poorly predicted by patterns of rainfall), with distinct dynamics separating rural and urban settings. We show that this variation has material consequences for the effective design of entomological surveillance programmes. Integrating these results with a previously published model of malaria transmission also highlights how this variation will influence the efficacy of malaria control efforts in parts of the Horn of Africa where the disease is currently (or has previously been) largely absent and underscores the need for rapid scaleup of entomological monitoring across the region.

## Methods

### Systematic Review of *Anopheles stephensi* Surveys

We collated references from published systematic reviews of literature relating to *An. stephensi*^16,35^, and updated these previous searches by searching *Web of Science* and *PubMed* from Jan.2017 to Sep.2020. We included all records containing temporally disaggregated adult mosquito catch data with monthly (or finer) temporal resolution spanning at least 10 months, that had not been conducted as part of vector control intervention trials and where at least 25 *An. stephensi* mosquitoes had been caught over the study period. A total of 36 references were collated containing 65 time-series with monthly catch data (no study presented data at a finer temporal resolution) from surveys carried out across Afghanistan, Djibouti, India, Iran, Myanmar and Pakistan. See **Supplementary Information** for further details and references therein.

### Clustering of Similar Time-Series & Random Forest Prediction of Cluster Membership

Following methodologies developed in previous work^35^, we fitted a Gaussian Process-based model to smooth these mosquito count time-series, using a Negative Binomial likelihood to account for overdispersion and a periodic kernel function to capture the repeating patterns often observed seasonally in mosquito populations. Model fitting was carried out within a Bayesian framework, using the probabilistic programming language STAN^36^. We then calculated summary statistics for each smoothed time-series to characterise their temporal properties (**Supplementary Information**), generating a set of parameters for each time-series that summarises their temporal properties. We then scaled and normalised each summary statistic to give a mean 0 and unit variance –a process necessary for the principal components analysis (PCA) we apply to identify a lower-dimensional representation of the structure present in the data amenable to visualisation.

Using *k*-means clustering, we identified clusters of time-series with similar temporal properties – the output of this process is a label for each time-series indicating which cluster (of time-series with similar temporal properties) each specific time-series was assigned to. For each study location, we extracted a suite of satellite-derived environmental variables (**Supplementary Table 2**) and used these variables alongside empirically calculated rainfall seasonality and average monthly catch as covariates within a random-forest based classification framework to predict cluster membership of each time-series. These models were fitted using the R package *Ranger*^37^ with 6-fold cross-validation utilised to optimise hyperparameters. Results are based on averaging the results of 25 iterations of cross-validation and model fitting and predictions made using out-of-bag estimates. There were significant imbalances in class size across clusters and so we carried out upsampling using the SMOTE^38^ algorithm. For results without upsampling see **Supplementary Information**.

### Probability of Detecting *Anopheles stephensi* With Different Surveillance Strategies

We explore the implications of seasonal variation in *An. stephensi* abundance on the probability of detecting the vector in entomological surveys using a theoretical sampling method with a defined amount of effort (such as a human landing catch conducted by a single volunteer for one night). We use a statistical framework (described further in **Supplementary Information**) that calculates the cumulative probability of detection from: i) an overall assumed *An. stephensi* annual biting rate (ABR, arbitrarily set to 20 for illustrative purposes here); ii) changes in vector density over the course of the year (from our collated time-series); and iii) various factors relating to timing of, and effort expended in, the entomological survey. Specifically, for each time-series, we identified the month with the highest rainfall, and the month in which vector density was highest (noting that these months were very rarely the same month). We then calculated the cumulative probability of *An. stephensi* detection using the framework, under a range of different surveillance strategies. Specifically, three strategies were simulated:

- **Vector-Peak Timed:** Starting the survey at the month with peak vector density (noting that in the absence of pre-existing detailed entomological information this is a hypothetical quantity designed to illustrate an approximate upper bound on the detection probability that could be achieved).
- **Rainfall-Peak Timed:** Starting the survey at the month with peak rainfall.
- **Random Month Timed:** The expected probability of detection achieved if the survey was started during a random month (calculated by simulating survey starting in each of the year’s 12 months and calculating the average cumulative probability).

In addition to varying the survey’s starting time, we also varied the amount of sampling effort (number of days sampled within each month) and overall duration of the survey (consecutive months sampled given a defined number of nights sampling per month). Note that the aim here is not to describe the exact probability of missing *An. stephensi* in any given entomological survey, as this will depend on a wide array of other, poorly defined and heterogeneous factors (e.g. type of catch methodology used, location etc). We also assume that the collection method is unbiased (i.e. not biased towards catching mosquitoes with particular resting or biting properties) which is also highly unlikely. Instead, the aim is to highlight how variation in seasonal dynamics can influence the nature of surveillance required to successfully detect a single *An. stephensi* (i.e. successfully establish presence), and the probability of detection should be viewed as a relative measure (i.e. viewed in relation to other sampling efforts and survey timings possible for surveys) and not an absolute value. Note that this framework assumes no seasonal variation in factors other than mosquito abundance (such as the ability of the sampling method to accurately record ABR) that might influence the probability of *An. stephensi* being caught by our theoretical sampling method.

### Modelling *Anopheles stephensi*-Driven Malaria Dynamics and Control

We integrated these vector abundance time-series into a published population-level model of *Plasmodium falciparum* malaria transmission and disease dynamics^39–41^ to explore the implications of *An. stephensi* seasonality on malaria control in settings in the Horn of Africa where malaria is currently largely absent (see **Supplementary Information** for full description of the modelling framework). We use the modelling framework to understand how variation in mosquito seasonality might influence the impact of IRS, a key vector control intervention. As the dynamics of *An. stephensi*’s establishment and influence on temporal trends in malaria transmission during its establishment remain unclear, we focused on the time-period immediately following establishment (when the disease is at equilibrium) and provide an illustrative example of how seasonality of *An. stephensi* driven malaria transmission could influence the effectiveness of IRS in a site with no pre-existing history of malaria control. For simplicity we assume that all transmission is due to *An. stephensi* and that IRS efficacy against this species is consistent with that observed against other species across the continent^42^. We simulate the impact of a single, illustrative IRS spraying campaign in a setting with an annual EIR of 1.5 (average malaria prevalence of 8-9%), timed either for optimal impact, randomly or alongside peak rainfall, and assume that 80% of the vector’s resting sites are successfully sprayed (noting the vector is thought to also rest in animal houses which are not typically sprayed in public-health campaigns). For further details, see **Supplementary Information**.

## Results

### Diversity in Temporal Dynamics Across the Collated *Anopheles stephensi* Time-Series

A total of 65 time-series from across Afghanistan, Djibouti, India, Iran, Myanmar and Pakistan were identified (**Fig.1A, Supp Fig.1)**. Substantial variation in the degree and timing of vector seasonality was observed, with the maximum percentage of annual vector density in any consecutive 4-month period (a proxy for degree of seasonality) ranging from 35-99% across the collated studies (average=62%). This contrasted with rainfall seasonality, where highly seasonal rainfall patterns were consistently observed across the locations the surveys had been carried out in (maximum percentage of annual rainfall in any consecutive 4-month period, mean=82, range 47-99%). We also observed a diverse range of temporal patterns, ranging from highly seasonal dynamics with a single seasonal peak, to bimodal population dynamics with two peaks within a single year, or more perennial patterns of abundance (**Fig.1B**).

**Figure 1:**
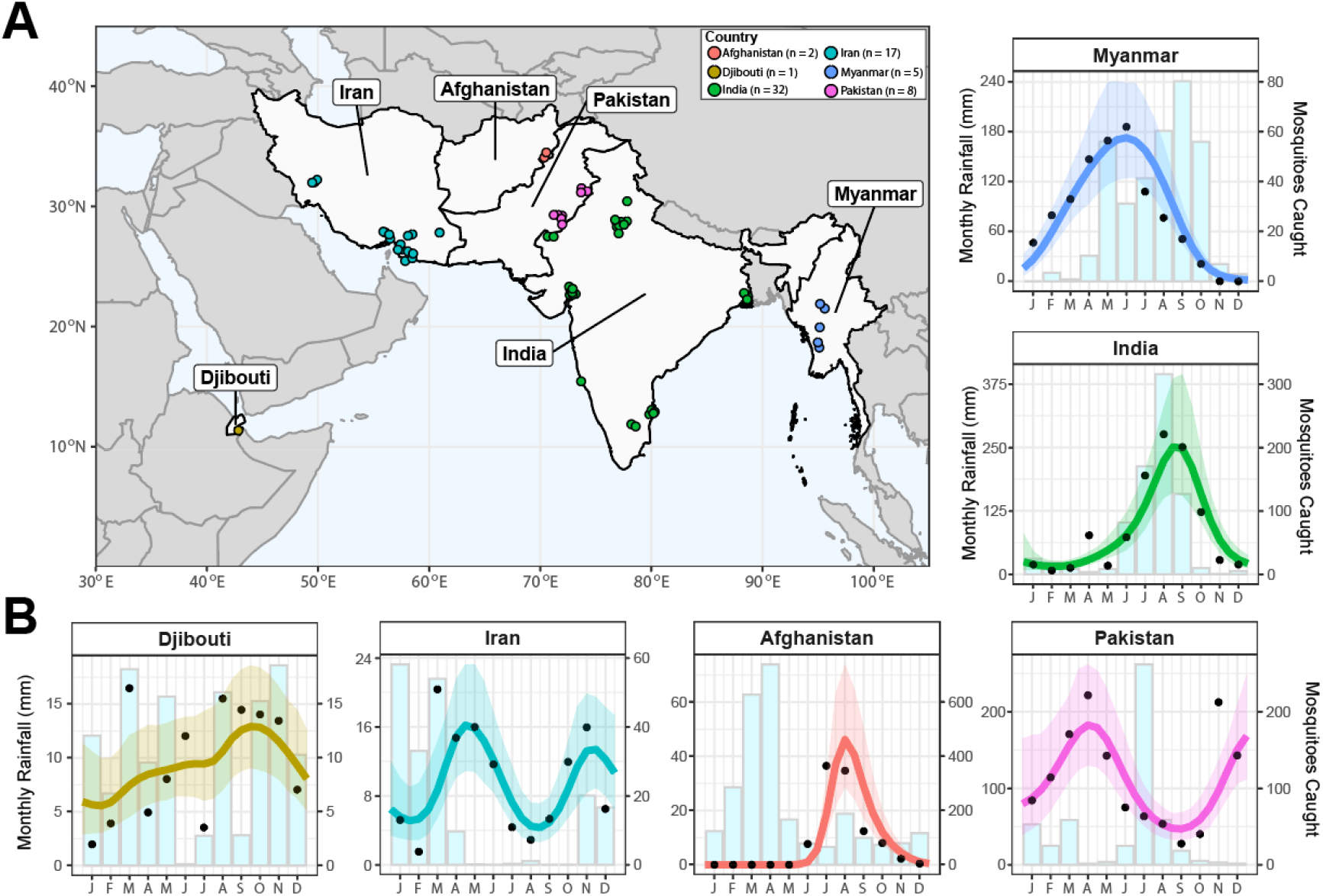
Sources and Locations of *Anopheles stephensi* Time-Series Data and Examples for Each Country. **(A)** Map of the geographical range over which time-series entomological collections have been carried out. Countries with studies are highlighted in light grey, and the locations of individual studies indicated by the individual points, coloured according to country (Afghanistan=red, Djibouti=yellow, India=green, Iran=turquoise, Myanmar=blue and Pakistan=pink). **(B)** Example *An. stephensi* time-series from each country, with the empirical monthly mosquito catch (black points), fitted gaussian process curves (mean=coloured line, ribbon=95% Bayesian Credible Interval) and monthly rainfall (matching sampling location and year of sampling) for each (light blue bars). The x-axis indicates the month of sampling, the y-axis either the monthly rainfall (left hand side y-axis) or number of vectors caught in each month (right hand side y-axis; note that the absolute number of mosquitoes caught between time-series are not comparable due to variable sampling effort). *n* indicates the number of time-series in each country.

### Statistical Characterisation and Clustering of Temporal Properties Highlights Distinct Archetypes

Summary statistics were calculated for each time-series to characterise their temporal properties (**Supp Fig.2**), followed by k-means clustering of the results to cluster the time-series into groups with similar temporal patterns. Our results highlight two distinct clusters of time-series, each characterised by distinct temporal patterns (**Fig.2B**). Cluster 1 time-series had single seasonal peaks and were more seasonal (average of 68% of annual vector density in the consecutive 4-month period with highest density) than Cluster 2 time-series, which had more perennial patterns of annual abundance (average 44% of annual vector density in the consecutive 4-month period with highest density) and contained several time-series with two peaks across the year. Despite differing significantly in mean vector abundance seasonality **(Fig.2C** and **2D**, p<0.001), there was no significant difference between Clusters in rainfall seasonality **(Fig.2D**, p=0.59). Seasonality of rainfall (defined as the highest proportion of total annual rainfall occurring in any consecutive 4 month period) across sampled locations was high (average 82% and 84% for Clusters 1 and 2 respectively) despite wide variation in vector abundance seasonality. Timing of peak rainfall relative to peak vector density significantly differed between clusters (**Supp Fig.3)**, with peak rainfall and vector abundance separated by <1 month on average for Cluster 1 compared to 2.2 months for Cluster 2. There was, however, considerable within-cluster variation in timing–within Cluster 1, timing of peak vector density relative to rainfall ranged from -5.8 months to +5.3 months (with 6 months the maximum gap that can occur within an annually repeating 12 month time-series, highlighting that the peaks in vector density relative to rainfall were found across the entirety of the year). We also explored varying the number of clusters specified in the k-means algorithm. Specifying 4 clusters resulted in further disaggregation of the 49 time-series in Cluster 1 into 3 separate clusters, each characterised by a single seasonal peak, but which differed in the timing of peak vector-density relative to peak rainfall (**Supp Fig.4**).

**Figure 2:**
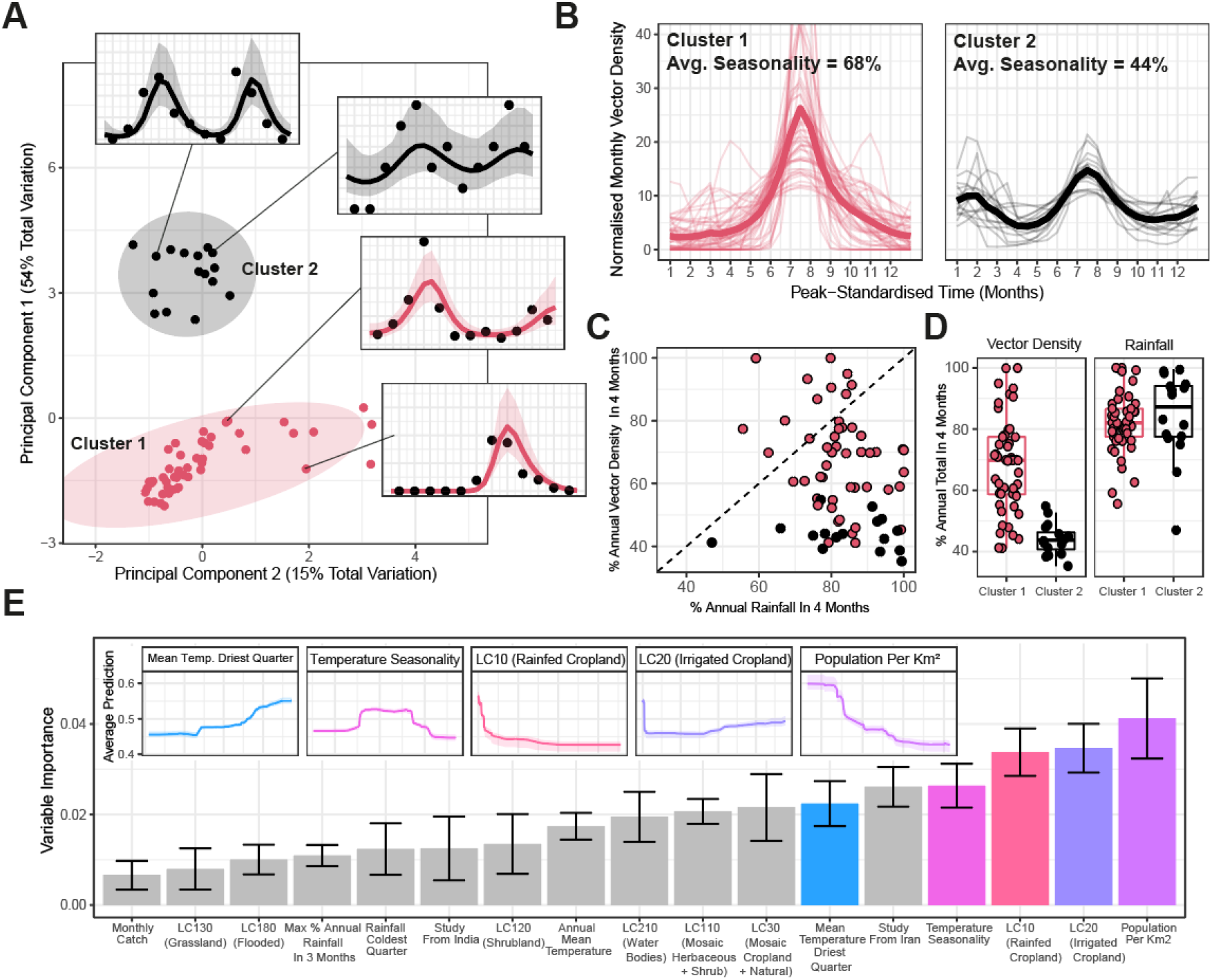
Characterisation and Clustering to Identify Time-Series with Similar Temporal Properties. **(A)** Results of principal components analysis (PCA) and k-means clustering for 2 clusters. Points on main figure indicate individual time-series, with point colour indicating cluster membership. Ellipsoids demarcate the 75^th^ quantile of the density associated with each cluster. Principal components 1 and 2 are plotted, together explaining 69% of the total variation in temporal properties across the time-series. **(B)** Time-series belonging to each cluster. Pale lines represent individual time-series, brighter line the mean of all the time-series belonging to that cluster – in all cases vector density is normalised to sum to 1 over the course of the year, and time-standardised so that the highest vector density for each time-series is arbitrarily set to occur at month 7. **(C)** Plot comparing the percentage of annual total mosquito catch and percentage of annual total rainfall occurring in any consecutive 4-month period for each time-series, coloured by cluster membership. **(D)** Boxplots show the percentage of annual total mosquito catch (left) and annual total rainfall (right) series occurring any in consecutive 4-month periods for each time-series. Rainfall data comes from the *CHIRPS* dataset^37^ and is specific to study location and time-period. Each point indicates an individual time-series. **(E)** Variable importance plot for each of the covariates included in the random forest model used to predict cluster membership– bar height indicates the mean variable importance across the 25 individual iterations of random forest fitting, with error bars representing the 95% confidence interval. Inset plots are the partial dependence plots for the top 5 most important variables in the model showing how the average prediction for Cluster 2 (y-axis, with higher values indicating an increased probability of Cluster 2 membership) varies with (normalised) variable value (x-axis).

### Random-Forest Modelling of Seasonal Dynamics Highlights Urbanicity as a Key Factor

We fitted a random forest-based classification framework to predict cluster membership (Cluster 1 or Cluster 2, as defined in **Fig.2A**). Due to the significant class size imbalance between Cluster 1 (n=49) and Cluster 2 (n=16), we up-sampled Cluster 2 data to generate balanced classes. Across 25 iterations of random forest model fitting, mean AUC was 0.89 (indicating good predictive performance) and the model was able to correctly classify Cluster 1 and Cluster 2 time-series equally well (83% and 85% accuracy respectively).

We calculated the relative importance of each variable to the model’s predictive ability **(Fig.2E)**. Patterns of land-use were strongly associated with different clusters–time-series from surveys in locations with lower population density (a proxy for rurality) more likely to belong to Cluster 2 (less seasonal), as were areas with a high proportion of land occupied by irrigated cropland. By contrast, a high proportion of land occupied by rainfed cropland was associated with Cluster 1 (more seasonal) dynamics. We also observed strong associations with temperature covariates, including the mean temperature of the driest quarter (where a high temperature was associated with Cluster 2), temperature seasonality (where a non-monotonically increasing relationship was observed, see **Fig.2E inset panels** and **Supp Fig.5** for all covariate response plots) and whether the study had been conducted in Iran (indicating potential spatial confounding). By contrast, rainfall seasonality was not an important predictor of temporal dynamics and was in the least 5 important variables. Examining the association between cluster membership and rurality/urbanicity (defined by study authors), there was indication of an association (chi-squared test, p=0.07), though this was not statistically significant at the 5% level. 88% (n=22/25) time-series from urban settings were assigned to Cluster 1, and only 12% (n=3/25) assigned to Cluster 2. 65% (n=24/37) of time-series from rural settings were assigned to Cluster 1, and 35% (n=13/37) to Cluster 2.

Model predictive performance and variable importance rankings were similar when no up-sampling was applied (AUC=0.81, **Supp Fig.6**), though predictive accuracy was highly unbalanced (Cluster 2 accuracy=50%, Cluster 1 accuracy=94%). Model performance and variable importance ordering remained similar when fitting the model and explicitly holding out a subset of the data to subsequently evaluate model performance (n=7 time-series, **Supp Fig.7**). Predictive power for seasonality (percentage of vector catch in any 4-month period) was more modest, although estimates were positively correlated (r=0.43, **Supp Fig.8**).

### Implications of Seasonal Dynamics for Entomological Surveillance of *Anopheles stephensi* across the Horn of Africa

We collated the same covariates for countries across the Horn of Africa and used the random forest model to predict cluster membership and potential temporal dynamics of *An. stephensi* across the region (**Fig.3A**). Our results highlight distinct geographical areas considered more likely to belong to Cluster 1 (more seasonal) and Cluster 2 (less seasonal), as well as areas of significant uncertainty. We next asked what consequences this seasonality might have on entomological surveillance of the vector, with a focus on how these seasonal dynamics might interact with features of surveillance programmes such as the timing and duration of entomological surveys. Across the collated temporal profiles, in a setting with an ABR of 20, surveys consisting of 3 months sampling and 3 sampling days per month that were timed to start at periods of peak *An. stephensi* density were on average 64% more likely to detect the vector compared to starting the survey at a random month of the year; and 57% more likely to successfully detect the vector compared to starting the survey in the month of peak rainfall **(Fig.3B)**. Timing entomological surveys to coincide with peaks in rainfall did not lead to a significant increase in the probability of successfully detecting *An. stephensi* (average 4% increase), suggesting that the timing of peak rainfall may be a poor measure for guiding entomological surveys searching for the vector. We next stratified these results by temporal Cluster **(Fig.3C)**. For Cluster 1 (and a survey lasting 3 months, with 3 days sampling per month) we observed differences in the cumulative probability of detection when comparing strategies which start surveys at the location’s rainfall peak, compared to starting them at peak *An. stephensi* abundance – on average, the latter strategy increased the cumulative probability of detection by 62% compared to a randomly timed survey, compared to only a 40% increase over random timing for Cluster 2.

**Figure 3:**
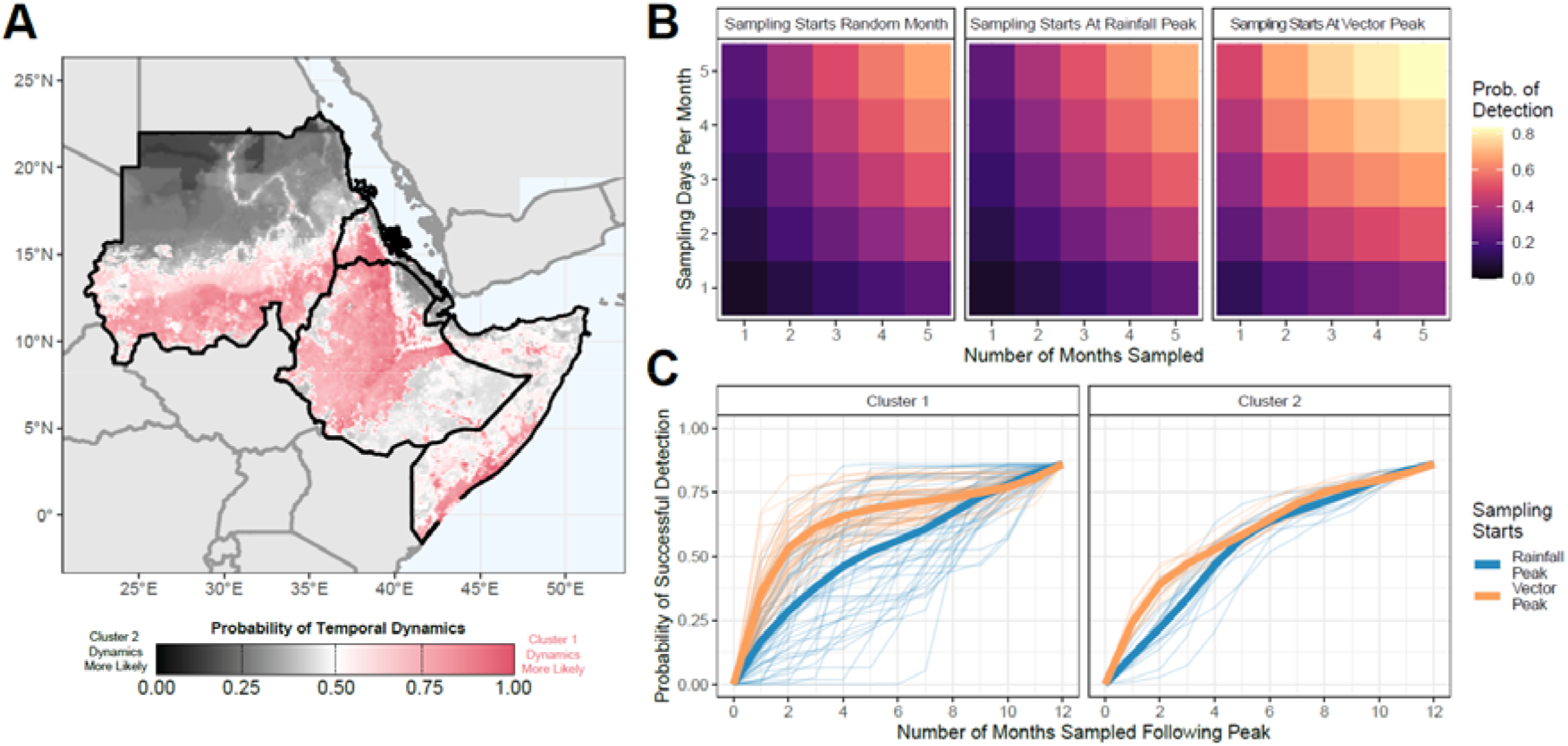
Possible Seasonal Dynamics of *Anopheles stephensi* Across the Horn of Africa, and Consequences for Entomological Surveillance and Monitoring. **(A)** Environmental covariates were collated across countries in the Horn of Africa where *An. stephensi* has been found, and the random forest classification model from **Fig 2E** used to predict potential temporal dynamics. Map shows the probability of temporal dynamics belonging to Cluster 1 (more seasonal), with pink corresponding to Cluster 1 dynamics being more likely and black indicating Cluster 2 dynamics (more perennial) are more likely, with white indicating both are equally likely. **(B)** For a setting with an annual biting rate of 20, the average probability (across all 65 collated *An. stephensi* temporal profiles) of detecting *An. stephensi* (where detection is defined as catching ≥1 mosquito) for a range of different sampling efforts (number of consecutive months sampled and number of sampling days in each month) in a setting with an annual biting rate of 20 bites per person. These results were generated for 3 different sampling strategies: i) with sampling starting at a random month in the year (left hand panel, subsequently averaged over all possible sampling start months in the year); ii) with sampling starting in the month of peak rainfall (centre panel); or iii) with sampling starting in the month of peak vector density (right hand panel). **(C)** For setting with an annual biting rate of 20 and a sampling effort of 3 days per month, the cumulative probability of *An. stephensi* detection as a function of the number of consecutive months sampled for each individual time-series, stratified by sampling strategy (starting at peak vector abundance=orange, at peak rainfall=blue) and Cluster. In both panels, pale, thin lines indicate the cumulative probability curve for a specific temporal profile, and thicker lines indicate the average for the specific sampling strategy.

### Modelling the Impact of *Anopheles stephensi* Seasonality On Vector Control Measures

Integrating the temporal profiles of *An. stephensi* abundance with a malaria transmission model, we explored how variation in temporal dynamics influences the impact of IRS (with two different insecticides, **Fig.4A**). Across the *An. stephensi* temporal profiles, optimal timing of IRS delivery resulted in an average of 47.6% reduction in annual malaria incidence in the 12 months following spraying for pirimithos methyl, and 28.9% for bendiocarb **(Fig.4B)**. These results represent 1.12x and 1.41x increases over the average impact achieved if the campaign is timed to a random month of the year. The extent to which optimal timing provided greater impact than random timing was dependent on the degree of seasonality and insecticide however–it increased with the degree of seasonality and was consistently larger for bendiocarb than pirimithos methyl (due to the latter’s longer duration and retention of residual activity following spraying). Timing the IRS campaign to occur when rainfall peaks did not significantly increase impact compared to timing the campaign to a random month (with less than a 2% average increase in impact for both pirimithos methyl **(Fig.4C)** and bendiocarb **(Fig.4D)**) and had significantly lower impact than optimally timed campaigns (39% and 15% lower impact for pirimithos methyl and bendiocarb respectively).

**Figure 4:**
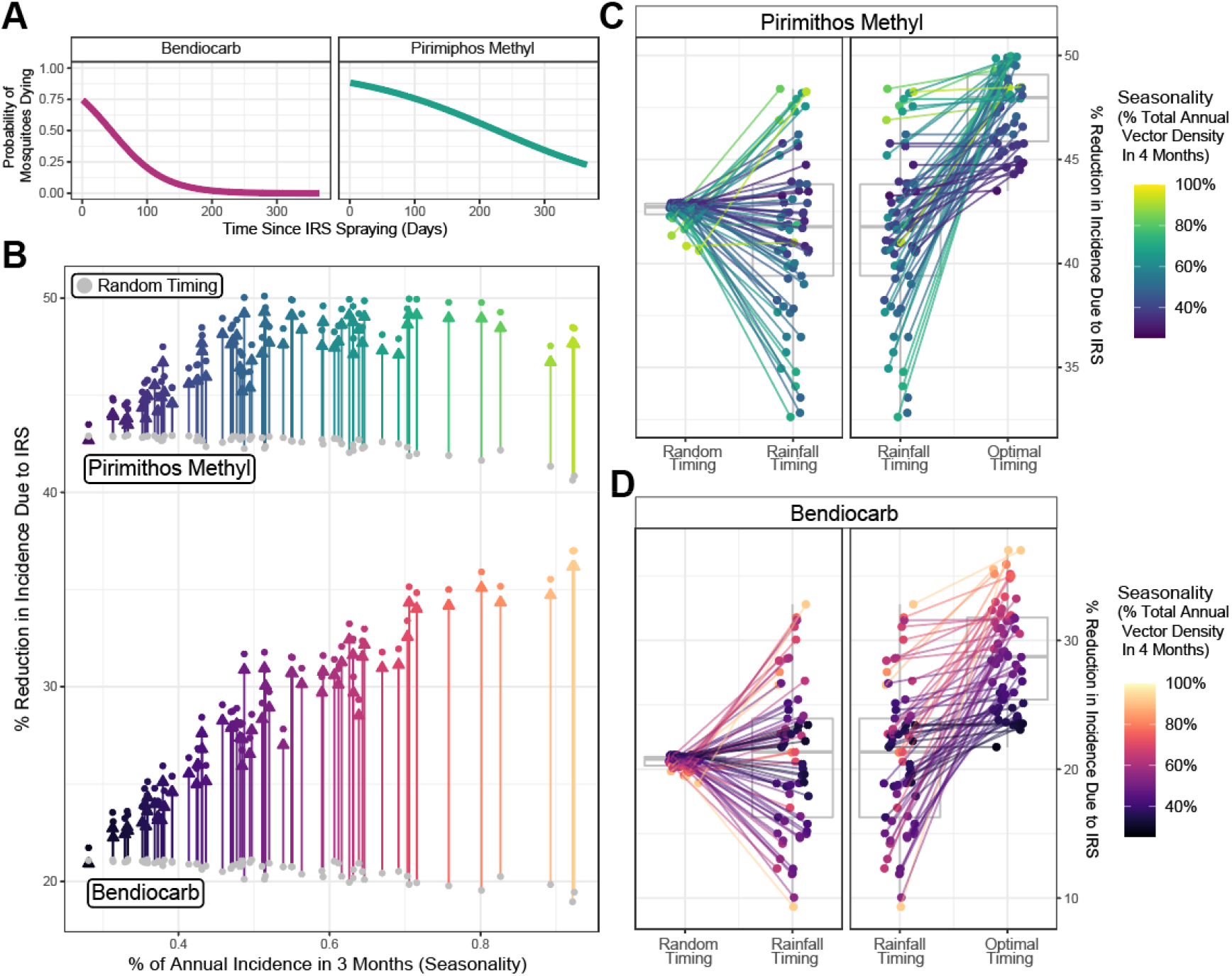
Modelling the Public-Health Impact of Indoor Residual Spraying and the Influence of *Anopheles stephensi* Seasonality. **(A)** Probability of mosquitoes dying upon exposure to each IRS compound in the time-period following spraying^42^ – pink indicates bendiocarb, turquoise indicates pirimiphos methyl. **(B)** For each temporal profile, the public health impact of annual IRS campaign with each insecticide according to the timing of the campaign. Points in grey correspond to the average reduction in incidence occurring from picking a random month to conduct the IRS campaign for each *An. stephensi* temporal profile, coloured points indicate the reduction in incidence arising from optimally timing the IRS campaign relative to the vector density peak for each *An. stephensi* temporal profile (and coloured arrows indicate the difference). The arrows and coloured points are coloured according to i) the insecticide used and ii) the degree of *An. stephensi* seasonality in each temporal profile (defined as the proportion of total annual abundance in any consecutive 4-month period). **(C)** For pirimithos methyl and for each *An. stephensi* temporal profile (coloured points) the percentage reduction in malaria incidence achieved if the IRS campaign is timed randomly, timed to start when rainfall is at its peak, or optimally timed based on peak vector density. Points correspond to specific *An. stephensi* temporal profiles and are coloured according to their degree of seasonality. Boxplot shows the minimum, first quartile, third quartile and maximum for the different individual projections. **(D)** As for **(C)**, but for the insecticide bendiocarb.

## Discussion

Invasion and establishment of *An. stephensi* across the Horn of Africa represents an urgent threat to malaria control in the region. Understanding the temporal profile of vector abundance of the species will inform effective deployment of surveillance, monitoring and control interventions aimed at mitigating this potential impact, particularly in urban settings where malaria has historically been largely absent or only minimally present. Collating data from across the vector’s endemic range, we identify broad diversity in the extent and nature of *An. stephensi* seasonal dynamics. This variation is associated with a wide array of ecological factors, including seasonal fluctuations in temperature and patterns of land use, including a potential role of urbanicity in shaping dynamics.

Our analyses identified population per km^2^ as the most important predictor of cluster membership, with high population density being strongly associated with Cluster 1 dynamics (more seasonal patterns of abundance). This potential disparity in temporal dynamics across rural and urban settings will likely have implications for both how resources aimed at surveillance control should be targeted to these different settings, and the public health impact of different control interventions. Our results suggest that urban *An. stephensi* populations are likely to display more seasonal dynamics, supporting the utility of temporally targeted interventions like short-lived IRS or LSM in these settings. The same is not necessarily true in rural settings, where shorter duration control interventions are likely to be impactful but may be less consistent in their effectiveness (without local surveys being conducted to establish the timing and extent of seasonality) due to the range of seasonal profiles observed, which included more perennial patterns of abundance. Implementing these measures and achieving sufficient intervention population coverage in urban settings is likely to present logistical challenges, given the historical absence of large-scale vector control campaigns from urban communities. If these barriers can be surmounted however, our results suggest such measures are likely to be impactful, though remaining uncertainty around the degree of endophily *An. stephensi* can display^43^ might necessitate alternative interventions to IRS that are not dependent on resting behaviour, such as LSM^34^.

Our results also suggest a limited role for rainfall in shaping the diverse temporal dynamics across the collated *An. stephensi* catch-data, contrary to results observed for other Africa malaria vector species (e.g. *An. gambiae*^44,45^). Specifically, that areas with highly seasonal rainfall may not have highly seasonal patterns of *An. stephensi* abundance. Instead, our analyses highlight an association between temperature and seasonal patterns of abundance with both temperature seasonality and the average temperature during the driest quarter being highly predictive of dynamics. This is consistent with previous work identifying temperature as a key driver of mosquito population dynamics, due to its impact on an array of mosquito life-history traits including biting rate, lifespan and fecundity (amongst several others)^46,47^. It should be stressed however that these covariates identified here are not necessarily predictive of absolute *An. stephensi* abundance in a region, but rather the seasonality in abundance. A much more detailed sampling strategy considering variability in the accuracy and biases of sampling methods and other geospatial methods will be needed to identify whether the vector has invaded a region.

The work also highlights the exceptionally limited amount of longitudinally collected entomological data from across *An. stephensi’s* current geographical range (including the Horn of Africa region) that currently exists. In highly seasonal settings there is a risk of erroneously concluding *An. stephensi*’s absence, particularly as the time of low vector catches may not coincide with times of low rainfall, as is frequently the case for other mosquitoes endemic to Africa. Longitudinal surveys enabling better description of these dynamics would therefore be useful in enabling subsequent refinement and timing of shorter surveys aimed at detecting presence only (whilst also providing additional information on temporal dynamics that can facilitate the effective targeting and timing of interventions such as IRS or LSM). Indeed, our results suggest that rainfall may provide a poor guide to timing of intervention campaigns in settings where *An. stephensi* is the dominant vector, underscoring the crucial role detailed entomological data collection and establishment of patterns empirically will play in optimising vector surveillance and disease control efforts.

There are several important limitations to the work presented here. Firstly, we assume that the inferred ecological relationships linking environmental features to temporal dynamics will translate from the vector’s historical range to the Horn of Africa. Indeed, our results highlight significant plasticity and variation in *An. stephensi*’s seasonal abundance depending on the setting, and therefore the extent to which our results will extrapolate to new settings remains unclear–making collection and analysis of longitudinal catch-data collected from the Horn of Africa an urgent research priority. Relatedly, due to the limited amount of data available and the wide geographical range over which the collated studies were conducted, we cannot rule out possible spatial confounding in shaping the inferred associations. Analysis of the distribution of locations stratified by rural/urban status and cluster assignment did not reveal obvious patterns of spatial confounding (**Supp Fig.9**), though the study being conducted from Iran was a high-ranking variable in the random forest model and it is possible some degree of spatial confounding is present. We were also unable to consider is the possibility of variation in temporal dynamics between *An. stephensi* forms. Identification of the *An. stephensi* form is challenging, requiring close visual examination^48^ or molecular methods^49^. Availability of this data was limited, and we lack the ability to disaggregate time-series by the specific form caught and hence preclude form as a confounder of some of the identified relationships linking environmental factors and temporal dynamics. Another limitation relates to our usage of mosquito abundance data, which is highly prone to biases driven by the collection method used. Whilst the analyses here use normalised mosquito counts (rather than raw abundance), trapping method bias might vary between season, which could affect the reliability of the seasonal patterns inferred, as well as introduce additional variation into catch data not captured here and which would influence the results describing the probability of detecting *An. stephensi* under different sampling effort and survey timing contexts.

We do not include insecticide resistance into our model of malaria transmission. Insecticide resistance is well-documented for *An. stephensi*^50–52^, and recent populations assayed in Ethiopia showed resistance to all four major insecticide classes^53,54^, suggesting that IRS might have a lower impact than suggested here. Relatedly, we do not consider uncertainty in *An. stephensi* bionomic properties (e.g timing of biting or whether resting occurs predominantly indoors or outdoors), which might vary by season and could further modulate the impact of interventions such as IRS where killing is mediated primarily through indoor resting following feeding. Variation in *An. stephensi*’s bionomic properties has previously been identified^55^, including a propensity for crepuscular biting and resting outside of houses compared to other *Anopheles* species dominant in sub-Saharan Africa^16,19,43^ that might render IRS less effective and necessitate consideration of other strategies such as LSM. Whilst the aim of this work is to illustrate how seasonality modulates (rather than precise estimates of) intervention impact, these considerations underscore the urgent need for a more detailed characterisation of *An. stephensi* across the Horn of Africa to quantify its bionomic properties and insecticide resistance profile more precisely in these settings, and identify the most effective control interventions to deploy.

Our work highlights significant variation in temporal dynamics across *An. stephensi* populations; variation that is shaped by distinct ecological factors, can markedly differ between urban and rural settings, and which has material consequences for the effectiveness of vector control interventions. Our work also highlights the need to better understand the vector’s dynamics in settings where it has newly established, and how these dynamics might differ to other *Anopheles* species also present and capable of malaria transmission. Indeed, the trajectory of *An. stephensi*’s establishment and subsequent dynamics in the Horn of Africa remains deeply unclear and the scarcity of published entomological studies from the region underscores the need for studies longitudinally surveying locations where *An. stephensi* has recently arrived. This will be important to understanding the patterns of seasonal variation the vector displays, and support optimising the delivery of malaria control interventions aiming to mitigate the impact of this invasive vector.

## Data Availability

All data collated as part of this study and the code required to reproduce these analyses can be found at the following link: https://github.com/cwhittaker1000/stephenseasonality.

## Acknowledgements and Funding Sources

CW is supported by Sir Henry Wellcome Postdoctoral Fellowship, Ref 224190/Z/21/Z. This research was funded in whole, or in part, by the Wellcome Trust (Ref 224190/Z/21/Z). For the purpose of Open Access, the author has applied a CC BY public copyright licence to any Author Accepted Manuscript version arising from this submission. SB & AG both acknowledge grant support from the Bill and Melinda Gates Foundation. TSC and AH both acknowledge Wellcome Trust (National Institute for Health Research–Wellcome Partnership for Global Health Research Collaborative Award, ‘Controlling emergent Anopheles stephensi in Ethiopia and Sudan (CEASE), Ref: 220870_Z_20_Z). The work is supported by the MRC Centre for Global Infectious Disease Analysis (reference MR/R015600/1) which is jointly funded by the UK Medical Research Council (MRC) and the UK Foreign, Commonwealth & Development Office (FCDO), under the MRC/FCDO Concordat agreement, the EDCTP2 programme supported by the European Union and Community Jameel. GCD acknowledges funding from the Royal Society.

## Author Contributions

CW, AH and TC conceived the study. ESS and SB contributed to the design of the study. CW carried out the systematic review. CW and SB developed the underlying statistical framework, with input on the analyses from AH, AG, TC, GCD, PGTW, PW and ESS. CW wrote the first draft manuscript, with all authors providing feedback and suggestions during manuscript drafting. All authors approved the final version of the manuscript.

