## Supplementary Information for "Seasonal Dynamics of *Anopheles stephensi* and its Implications for Mosquito Detection and Emergent Malaria Control in the Horn of Africa"

**Contents**

**Supplementary Information 1:** Description of Systematic Review, Data Extraction and Initial Processing

**Supplementary Information 2:** Description of Statistical Methodologies Utilised

**Supplementary Information 3:** Additional Figures and Results

**References**

**Outline of Document**

In this supplementary document we outline the methods and data used to explore and analyse the patterns and drivers of *Anopheles stephensi* population dynamics across South Asia and the Middle East. In **Supplementary Information 1**, we present an overview of the systematic search strategy used to collate the references containing the extracted and analysed data, as well as details about the initial pre-processing steps applied to said data. In **Supplementary Information 2**, we describe the statistical methodologies used to process and analyse this extracted data. The output of these analyses forms the basis for the results presented in the main text. Finally, in **Supplementary Information 3**, we present a number of additional figures and tables to support the work detailed in the main text.

**Supplementary Information 1: Description of Systematic Review: Data Extraction and Initial Pre-Processing**

**Systematic Review: Search Procedure and Record Screening**

We collated references from two previously published systematic reviews of literature relating to *Anopheles stephensi* (focusing on its presence/absence across a wide geographical range^1^ and its seasonal dynamics in India^2^ respectively), and updated these previous searches (both conducted in 2017) by searching *Web of Science* and *PubMed* databases from January 2017 for further relevant references containing temporally disaggregated *Anopheles stephensi* catch data. Key words for this search were:

(((anophel*) AND ((India) OR (BURMA) OR (MYANMAR) OR (BANGLADESH) OR (THAILAND) OR (ISLAMIC REPUBLIC OF IRAN) OR (ETHIOPIA) OR (DJIBOUTI) OR (SUDAN))) AND (("2017"[Date - Publication] : "3000"[Date - Publication])) OR ((anophel*) AND ((Pakistan) OR (Iran) OR (Afghanistan)) AND (("1990"[Date - Publication] : "3000"[Date - Publication]))

with references for Pakistan, Iran and Afghanistan searched for over an extended time-period (i.e. date range of 1990-2020 rather than 2017-2020) to ensure completeness of the collated references, and fill in countries not included during previous reviews. Our searches identified a total of 926 records, which were screened according to the following Inclusion/Exclusion criteria:

**Inclusion Criteria:**

- Reference contains temporally disaggregated adult mosquito catch data for *An. stephensi*, at a temporal resolution of monthly or finer.
- The time-period spanned by the survey must be at least 10 consecutive months in duration and have caught at least a total of 25 *An. stephensi* over the period for which catches were being carried out.

**Exclusion Criteria:**

- Mosquito catch data is not temporally disaggregated to a sufficient extent (e.g. catches were done yearly or seasonally rather than monthly).
- Mosquito catch data was collected as part of a trial assessing a vector control intervention (which would perturb the natural dynamics of the vector, rendering the data unrepresentative of the population dynamics in the absence of control).
- The reference only contained information on immature/larval mosquito life cycle stages rather than mature adults.
- The reference contained insufficient information to geolocate the area in which the study was conducted to at least the administrative unit 2 level.

Overall, a total of 34 references were collated containing 65 time-series from catch surveys carried out in distinct locations from across Afghanistan (n=2), Djibouti (n=1), India (n=32), Iran (n=17), Myanmar (n=5) and Pakistan (n=8). These were further supplemented with 2 references (from Pakistan and India respectively, yielding a total of 3 time-series) collated as part of a (currently unpublished) systematic review of the bionomics of secondary malaria (i.e., non-dominant) vectors across South Asia, yielding a total of 65 time-series from these 34 references. The next section describes in further detail about extraction and collation of the data associated with each study.

**Systematic Review: Data Extraction, Collation and Initial Processing**

**Entomological Data Extraction**

For each reference, we extracted all relevant entomological catch data provided that pertained specifically to *An. stephensi*. Where data were presented in a table, data was copied directly from the table. Where the data were in a graph, data were extracted using the DataThief^TM^ software. This yielded a total of 65 time series of monthly mosquito catch data (no reference presented data at a finer temporal resolution), ranging in length from 10 – 60 months, with a mean time-period of 15.6 months and a median time-period of 12 months, a mean catch size of 758 and a median catch size of 289.

**Supplementary Table 1: Number of time series collated according to method of mosquito collection.**

|  | Landing Catch | Resting Collections | Pit Collections | Light Traps | Pyrethrum Spray Catch |
| --- | --- | --- | --- | --- | --- |
| # Time-Series | 3 | 33 | 2 | 4 | 14 |

Of the collated studies, the majority sampled mosquitoes via resting collections (n=33), though there was significant variation between surveys as to where mosquitoes had been sampled (e.g. human dwellings or cattlesheds), when sampling had been carried out (daytime, night-time or overnight) and for the small number of landing catch studies collated (n=3), which bait had been used (cattle or humans). Of the 65 collated time-series, 56 presented results arising from a survey carried out using 1 catch methodology (described in **Supplementary Table 1** above). 9 time-series represented results which presented the total number of *Anopheles stephensi* mosquitoes caught across all methods of collection and could not be disaggregated by catch-type. They have not been counted in **Supplementary Table 1** above.

The primary focus of these analyses was to characterise annual and seasonal patterns of variation in *An. stephensi* abundance. Given this, and also that variations in time-series length are a factor known to affect their statistical properties^3^ (and therefore limit the comparability of the time series gathered and analysed here), all time-series were standardised to be 12 months in length. For time series containing more than 12 time points (i.e. time series that spanned longer than a single year), we averaged the recorded catches for a given month. For time-series containing less than 12 months of data, this was not carried out. Where the study has been initiated in a month other than January, and concluded in a month other than December, the recorded counts were reordered to yield a complete time series running from January to December (and then subsequently adjusted so that the month of peak vector density is arbitrarily set to month 7 when plotting the time-series, to enable graphical comparability, see **Fig.2A**).

The results presented in the collated references were frequently presented in the form standardised by sampling effort, such as Man-Hour Density (MHD). They do not therefore represent the total number of mosquitoes caught each month (required for the statistical framework utilised to characterise temporal properties) and therefore, where information on sampling effort was present (e.g. number of hours spent sampling/catching *An. stephensi*, number of households or cattlesheds searched, number of trap nights etc), we used this information to convert MHD back to the raw counts. In the small number of instances where there was variable sampling effort across the time series (which would bias the conversion away from the underlying population abundance), we conservatively used the lowest sampling effort recorded across the time series in the conversion. Together, this allowed us to produce an estimate of the number of mosquitoes sampled (a raw count, based on equal sampling effort across the time series). See **Supplementary Data: *Extracted Entomological Data*** for more information about how each time-series was processed).

**Study Geolocation and Environmental Covariate Extraction**

For each study where geolocation was possible, we recorded the location at both the administrative unit 1 and 2 level, based on information provided in the reference. A number of the references identified in our review had previously been utilised as part of previous reviews^1,2^ – where this data was available, these descriptions of study location were used. For each location, we then extracted a suite of satellite-derived environmental covariates. These environmental covariates consist of raster layers spanning all countries in which studies had been conducted in (i.e. Afghanistan, Djibouti, India, Iran, Myanmar and Pakistan) at a 2.5 arc-minute (~5km by 5km, depending on the exact location and distance from the equator) spatial resolution. The covariates utilised here were initially selected from a set of 19 derived from the *BioClimatic* variables (a suite of biological relevant covariates defined from monthly rainfall and temperature satellite data^4^, making the strong assumption that these variables, which represent location averages over the period 1970-2000, adequately describe the climactic factors present in the periods spanned by our studies, which were predominantly conducted after 2000) as well as measures of landcover and urbanicity^5^, population density^6,7^ and enhanced vegetation index^8,9^. This provided a total of 43 covariates, many of which were highly correlated with one another. To reduce the degree of this multicollinearity, we generated a reduced subset of covariates using tools available in the *tidymodels* collection of R packages^10^ that aim to minimise the Spearman rank correlation coefficients between retained covariates, and also exclude covariates where there is minimal variation for that covariate across the full dataset, leaving **19** covariates in total. In addition to the environmental covariates described above, for each of the administrative units a survey had been carried out in, we also collated daily rainfall estimates for the time-period the survey had been conducted in, using the *“The Climate Hazards Group Infrared Precipitation With Stations”* (CHIRPS) dataset^11^. These data were aggregated up to the same temporal resolution as the *An. stephensi* catch data (i.e. monthly). These rainfall data were used to calculate the cross-correlation coefficient between mosquito catches and rainfall.

**Supplementary Table 2: The Complete Suite of Covariates Collated and Subsequently Reduced for Modelling and Prediction of Seasonal Population Dynamics**

| **#** | **Variable** | **Temporal Resolution** | **Source** |
| --- | --- | --- | --- |
| 1 | BioClimatic - Annual Mean Temperature | Annual Average, 1970 - 2000 | <https://www.worldclim.org/bioclim> |
| 2 | BioClimatic - Mean Diurnal Range | Annual Average, 1970 - 2000 | <https://www.worldclim.org/bioclim> |
| 3 | BioClimatic - Isothermality | Annual Average, 1970 - 2000 | <https://www.worldclim.org/bioclim> |
| 4 | BioClimatic - Temperature Seasonality | Annual Average, 1970 - 2000 | <https://www.worldclim.org/bioclim> |
| 5 | BioClimatic - Max Temperature of Warmest Month | Annual Average, 1970 - 2000 | <https://www.worldclim.org/bioclim> |
| 6 | BioClimatic - Min Temperature of Coldest Month | Annual Average, 1970 - 2000 | <https://www.worldclim.org/bioclim> |
| 7 | BioClimatic - Temperature Annual Range | Annual Average, 1970 - 2000 | <https://www.worldclim.org/bioclim> |
| 8 | BioClimatic - Mean Temperature of Wettest Quarter | Annual Average, 1970 - 2000 | <https://www.worldclim.org/bioclim> |
| 9 | BioClimatic - Mean Temperature of Driest Quarter | Annual Average, 1970 - 2000 | <https://www.worldclim.org/bioclim> |
| 10 | BioClimatic - Mean Temperature of Warmest Quarter | Annual Average, 1970 - 2000 | <https://www.worldclim.org/bioclim> |
| 11 | BioClimatic - Mean Temperature of Coldest Quarter | Annual Average, 1970 - 2000 | <https://www.worldclim.org/bioclim> |
| 12 | BioClimatic - Annual Precipitation | Annual Average, 1970 - 2000 | <https://www.worldclim.org/bioclim> |
| 13 | BioClimatic - Precipitation of Wettest Month | Annual Average, 1970 - 2000 | <https://www.worldclim.org/bioclim> |
| 14 | BioClimatic - Precipitation of Driest Month | Annual Average, 1970 - 2000 | <https://www.worldclim.org/bioclim> |
| 15 | BioClimatic - Precipitation Annual Coefficient of Variation | Annual Average, 1970 - 2000 | <https://www.worldclim.org/bioclim> |
| 16 | BioClimatic - Precipitation of Wettest Quarter | Annual Average, 1970 - 2000 | <https://www.worldclim.org/bioclim> |
| 17 | BioClimatic - Precipitation of Driest Quarter | Annual Average, 1970 - 2000 | <https://www.worldclim.org/bioclim> |
| 18 | BioClimatic - Precipitation of Warmest Quarter | Annual Average, 1970 - 2000 | <https://www.worldclim.org/bioclim> |
| 19 | BioClimatic - Precipitation of Coldest Quarter | Annual Average, 1970 - 2000 | <https://www.worldclim.org/bioclim> |
| 20 | Population Density | 1992-2020, using the closest year value to the year of the study | <http://www.worldpop.org.uk> |
| 21 | Enhanced Vegetation Index | 1992-2020, using the closest year value to the year of the study | <https://modis.gsfc.nasa.gov/data/dataprod/mod13.php> |
| 22 | Landcover | 1992-2020, using the closest year value to the year of the study | <https://maps.elie.ucl.ac.be/CCI/viewer/index.php> |
| 23 | Average Monthly Catch | Calculated empirically for each time-series | NA |
| 24 | Maximum proportion of total annual rainfall in any consecutive 4 month period | Calculated empirically for each location and time-series | NA |
| 25 | Country survey had been carried out in | Calculated empirically for each time-series (grouped into “India”, “Iran” and “other”). | NA |

**Note:** There are **43** covariates total here, as Landcover contains **19** distinct covariates (each describing the proportion of cover attributable to a particular landcover class in a given area).

**Note:** All WorldClim data is from Version 2 of the datasets.

**Supplementary Information 2: Description of Statistical Methodologies**

**Negative Binomial Gaussian Process – Fitting and Inference:**

In-line with previously work modelling the seasonal dynamics of different *Anopheline* mosquito species from across India^2^, we utilise a flexible Gaussian Process modelling framework to temporally interpolate between the monthly-catch datapoints and smooth the raw, noisy and overdispersed catch data. Gaussian processes specify a distribution over functions such that any finite set of function values ${f(x}_{1}), {f(x}_{2}), \ldots{f(x}_{N})$ have a joint Gaussian distribution^12^. The Gaussian process is entirely specified by its mean function:

$$E\left[ f\left( x \right) \right]= \mu(x)$$

and by its covariance function:

$$Cov[f\left( x \right), f\left( x^{'} \right)]= k(x, x^{'})$$

The covariance function is also known as the kernel and defines, based on the Euclidean distance between any two points, their covariance (and thus the covariance matrix of the Gaussian Process when all pairwise combinations of points are considered). Many different forms of the kernel are possible that each encode different prior information about how we expect two datapoints ($x$ and $x^{'}$ in this instance) to be similar, and the distance over which we expect this similarity to persist. Given that mosquito population dynamics are typically characterised by seasonally repeating patterns occurring either, a periodic kernel function was used to define the covariance between pairs of points:

$$k\left( x, x^{'} \right)=\alpha^{2}exp\left( -\frac{2}{l^{2}}{sin}^{2}\left( \frac{\pi\left| x-x^{'} \right|}{p} \right) \right)$$

where $p$ represents the period over which we would expect points to show similar dynamics (i.e. a period of twelve would imply we expect points separated by 12 months to be most similar), $\alpha$ specifies the magnitude of the covariance, and $l$ represents a lengthscale parameter further constraining the extent to which two values separated by a given time can co-vary.

Bayesian inference and fitting of normal Gaussian Processes typically follow this hierarchical formulation:

$$\theta\sim\pi(\theta)$$

$$f \sim GP(0, K_{\theta}(x))$$

$$y_{i} \sim MVN\left( {f(x}_{i}), \sigma^{2} \right) \forall i \in\{1, \ldots, N\}$$

where $\theta$ represents a vector of hyperparameters involved in defining the kernel’s properties, $f$ is a distribution of functions from a zero-mean Gaussian Process with covariance function $K_{\theta}$, $f$(x) are function evaluations at times $x$, and $y$ the observed data. We modify this structure to account for specific characteristics of the mosquito data being utilised – specifically that the data are integer counts, that mosquito catch data is rarely normally distributed and frequently displays high levels of overdispersion (a common property of biological systems generally). We therefore adapted the above framework to accommodate a Negative Binomial likelihood, leading to the following inferential framework:

$p,\alpha,l \sim\pi( p,\alpha,l )$

$$\boldsymbol{f}\sim GP(0, K_{\theta}(x))$$

$$where: k\left( x, x^{'} \right)=\alpha^{2}exp\left( -\frac{2}{l^{2}}{sin}^{2}\left( \frac{\pi\left| x-x^{'} \right|}{p} \right) \right)$$

$$y_{i} \sim Negative Binomial\left( e^{{f(x}_{i})}, \sigma\right) \forall i \in\{1, \ldots, N\}$$

where $e^{f(x)}$ is used to reflect the fact that we use a log link between the observed counts and the underlying latent process reflecting the population dynamics, and $\sigma$ represents the overdispersion parameter of the Negative Binomial distribution.

**Prior Specification**

Per previous work^2^, prior distributions for the estimated parameters were defined as follows:

$$l \sim Normal(2, 1^{2})$$

$$\alpha\sim Half-Normal(0, \sqrt{SD (y)})$$

$$p \sim Normal(12, 4^{2})$$

$$\sigma\sim Half-Normal(0, 8^{2})$$

Weakly informative priors were set on the scaling factor $\alpha$, the period, $p$, and the overdispersion parameter, $\sigma$. The prior for the kernel period ($p$) was centred on $12$ (a value of the period that would represent annual variation being the dominant temporal modality) to reflect our prior belief that observed variation in mosquito abundance is likely to cycle annually. However, recognising that other temporal patterns of fluctuating abundance are possible, we placed a large standard deviation on $p$ to allow the model to accommodate instances of bimodality or periods operating across timescales longer than a year. We placed lower and upper bounds on $p$ at 4 and 18 months respectively, to avoid identifiability issues arising from the lack of data at temporal resolutions below and above these bounds.

**Model Fitting and Parameter Inference**

This Negative Binomial Gaussian Process were fitted using a Bayesian framework implemented in STAN, a probabilistic programming language for statistical inference written in C++ that employs the No-U-Turn sampler, a variant of the gradient-based Hamilton Monte Carlo algorithm for inference^13^. For each time-series, 2 chains of 20,000 iterations were run for purposes of model fitting and parameter inference. Half of each chain’s iterations were discarded as burn-in/the adaptive phase of the sampling, leaving a total of 20,000 iterations available for inference. Measures of MCMC convergence such as the Gelman-Rubin statistic were monitored in all cases and were all consistently < 1.02.

**Fitted Time Series Normalisation and Von Mises Distribution Fitting**

After having fitted and smoothed the mosquito catch time-series, we normalised each in the following way:

$$p_{i}= \frac{y_{i}}{\sum y_{i}}$$

where $p_{i}$ is the proportion of the annual catch recorded at timepoint $i$. This was done in order to establish comparability across the time series (which varied substantially in the absolute numbers of *Anopheles stephensi* caught). We then further characterised the periodic properties of these time series by fitting Von Mises distribution to the time-series. The Von-Mises distribution is a continuous probability distribution that exists on the circle, with range $0$ to $2\pi$. It is the circular analogue of the normal distribution (which exists on the line), with the probability density function for the angle $x$ given by:

$$f\left( x | \mu, \kappa\right)= \frac{e^{\kappa cos(x-\mu)}}{2\pi I_{0}(\kappa)}$$

where $I_{0}(\kappa)$ is the modified Bessel function of order 0, the parameter $\mu$ is a measure of location (analogous to the mean of the normal distribution, describing where on the circle the distribution is clustered around) and $\kappa$describes the concentration of density around $\mu$ (and thus its inverse is a measure of dispersion, analogous to $\sigma^{2}$ for the normal distribution. We fitted two sets of Von Mises densities to the normalised time series, the first containing a single component:

$$f\left( x | \mu_{1}, \kappa_{1} \right)=f_{1}\left( x | \mu_{1}, \kappa_{1} \right)$$

and another with two-components, formulated as:

$$f\left( x | \mu_{1}, \kappa_{1}, \mu_{2}, \kappa_{2}, w \right)=\omega f_{1}\left( x | \mu_{1}, \kappa_{1} \right)+(1-\omega)f_{2}\left( x | \mu_{2}, \kappa_{2} \right)$$

where $x$ represents the normalised mosquito count formulated as a random variable on the circle (i.e. $x=\frac{2\pi p_{i}}{12}$). Fitting was undertaken using the *optim* function in R, with the root mean squared error as the loss function. The outputs from this fitting were then included in the process generating aggregate summaries of the temporal properties of the time-series, a process described in further detail below.

**Time Series Characterisation and Analysis**

To characterise the temporal properties of each time-series, we calculated a series of summary statistics for each, drawing on previous work carried out exploring the empirical structure of time series^12^. In doing this, we can make explicit comparisons between time-series about key aspects of their temporal properties (e.g., the degree or timing of seasonality), and in doing so, identify time-series with similar statistical and temporal properties. These summary statistics were the following:

1. **Periodic Kernel Median:** Fitting the Negative Binomial Gaussian Process with a periodic kernel allowed inference of the period, $p$, providing us with an estimate of the frequency of repeating patterns in the monthly abundance of mosquitoes. An estimate of $p$ was calculated for each fitted time series, with the median value of $p$ across the X HMC iterations for each time-series used here
2. **Kullback-Leibler Divergence:** Also known as the relative entropy, the Kullback-Liebler divergence represents a measure of how different one probability distribution is from a second probability distribution (where a value of 0 indicates that the two distributions are identical). It is specified in the following manner:

$$E_{i}= p_{i}{log}_{2}\left( \frac{p_{i}}{q_{i}} \right)$$

$$E=\sum_{i=1}^{12} p_{i}{log}_{2}\left( \frac{p_{i}}{q_{i}} \right)$$

where $p_{i}$ is the average value of the normalised time series for month $i$, and $q_{i}$ = 1/12 for $i=1,\ldots, 12$. This operation therefore measures the deviation of a normalised time series from a uniform distribution, in doing so, informing about the extent to which a seasonal peak (or peaks) is present in the time series.

3. **Time Difference Between Vector Density Peak and Rainfall Peak Timings:** The time difference between the highest recorded vector density and the highest recorded rainfall for that year.
4. **Proportion of Points Greater Than 1.65x the Mean:** For each fitted, normalised time series, the proportion of points greater than 1.65x the time-series’ mean was calculated, informing the degree and width of any seasonal peaks.
5. **Number of Peaks:** Estimates of the parameters governing the fitted two component Von Mises distribution were used to infer the number of peaks in each time series. Specifically, and in-keeping with previous work^2^, a time series was deemed to possess one peak if the value of the Von Mises component weighting was either < 0.3 or > 0.7 and the difference in means was < $\frac{2\pi}{3}$ or > $\frac{4\pi}{3}$ , indicating that the majority of the density could be attributed to one of the two components, and that the two means identified during the fitting were temporally close to one another. Otherwise, a time series was judged to possess two peaks.
6. **Von Mises 1 Component Mean:** If a 1 component Von Mises distribution was preferred, then the Von Mises mean corresponding to the maximum likelihood predicted value was used. If the 2 component Von Mises distribution was preferred, the value for this operation for that particular time series is set to -5.
7. **Von Mises Two Component Weight:** Estimates of the weight parameter governing the two component Von Mises distribution were also used to infer the bimodality of the time series. The weight specifies the proportion of each component that is used to fit the time series and thus a very high (or very low weight) indicates the dominance of a single component and the comparatively small contribution of the other.
8. **Maximum Percentage of Total Annual Catch In Any 3 Month Period:** In-keeping with previous, operationally aligned estimates of malaria seasonality^14^, we calculated using a sliding 3-month window the maximum percentage of the total annual catch that was caught in any 3 month period.

**Principal Components Analysis and Clustering**

Principal Components Analysis (PCA) is a statistical procedure that utilises an orthogonal transformation to convert a set of correlated variables (in this case the outputs of the 7 mathematical operations described above for each of the time series) into a set of linearly uncorrelated variables (known as the “principal components”). In doing so, this allows us to summarise this set of variables with a smaller number of representative variables that together explain the majority of the variability in the variables. Reducing the dimensionality of the dataset in this way facilitates visualisation of time series properties (as defined by the mathematical operations) as well as clustering of the time series into groups which share similar properties (clustering algorithms typically perform poorly in high dimensional settings, necessitating the use of PCA as described here). Clustering was then undertaken using the k-means clustering algorithm, using the first four PCA components that together described 85% of the total variation present in the data.

**Random Forest Modelling and Prediction of Seasonality**

Random Forests are a machine learning, ensemble-based method that work by constructing a collection of decision trees that together explain the results (where results are either a continuous outcome variable in the regression context, or a binary indicator in the classification context)^15^. The outputs of these decision trees are subsequently aggregated in a statistically principled and coherent way to produce a “forest” (or ensemble) of trees that together produce predictions for comparison with data. They have previous been shown to provide significant improvements in accuracy over traditional linear regression based approaches, particularly in contexts where non-linear relationships or interactions between covariates are likely present and to be relevant to prediction of an outcome^16^.

We used a Random Forest based approach to either 1) classify time-series cluster membership (i.e. predict whether a time-series belonged to either Cluster 1 or Cluster 2, as defined via the PCA and k-means clustering analysis described above); or 2) predict *An. stephensi* time-series seasonality (defined as the percentage of total annual vector density in any continuous 3-month period). These models were fitted using the software package *Ranger*^17^, implemented in the *tidymodels* framework for R^10^, with 6-fold cross-validation utilised to optimise hyperparameter combinations; presented results are based on averaging the results of 25 separate iterations of cross-validation and model fitting (to account for stochasticity in model fitting), and any predictions made using out-of-bag model estimates in all instances. Due to significant imbalances in class size across the time-series clusters (49 time-series in Cluster 1 compared to only 16 time-series in Cluster 2, we carried out upsampling using the SMOTE (synthetic minority over-sampling technique^18^) algorithm. We also carried out model fitting without this upsampling, the results of which are presented in **Supp Fig. 6**.

In all instances, out-of-sample predictive accuracy was assessed using 6-fold cross-validation (CV) and used to optimise the hyperparameters associated with the Random Forest method algorithm. Random Forest models were fitted to the training dataset (i.e. the full dataset minus one of the CV folds) and then model accuracy assessed on the remaining fold of data not included in model training. In the case of the cluster classification example, the metric used to evaluate model performance was the area under the curve (AUC). In the case of the regression prediction of seasonality, the metric used to evaluate model performance was the root mean squared error (RMSE). The Random Forest hyperparameters providing the best out-of-sample AUC/RMSE were then selected, and a final Random Forest model then fitted on the full set of data available. Predictive accuracy (assessed via AUC/RMSE) was then calculated for the entire dataset by using out-of-bag predictions for each sample i.e. predictions on each training sample using only the trees that did not have that training sample in their bootstrap sample. We also calculated both permutation variable importance and generated partial dependency plots^19^ for each model to assess the contribution of specific, individual environmental covariates to whether a time-series had a single seasonal peak or not. Together these methods allow evaluation of the importance of each included covariate to model predictive accuracy, and in turn, allows us to “rank” covariates according to their contribution to the predictive performance of the model. This entire process was repeated 25 times in order to average over the stochasticity and variation inherent in the Random Forest fitting process.

We also carried out an additional sensitivity analysis where a set of the available data (n=12 time-series) was held-out at the onset, and the random forest model trained (using 6-fold cross-validation) on the remaining available data (n=53 time-series total, with 43 time-series used in model fitting and 10 time-series used for performance evaluation in each of the cross-validation folds). Optimal hyperparameters were selected in the same way as described above, and then a final model fitted to the full, non-held out data (n=53 time-series), and model predictive accuracy assessed by evaluating performance on the held-out data (n=12 time-series).

**Probability of Detecting *Anopheles stephensi* With Different Surveillance and Monitoring Strategies**

We explore the implications of seasonal variation in *An. stephensi* abundance on the probability of detecting the vector in entomological surveillance and monitoring using human landing catches. Note that what follows below assumes there is no seasonal variation in factors other than mosquito abundance (such as resting preferences) that might influence the probability of *An. stephensi* being caught in a human landing catch. In the absence of estimates of overall mosquito population size, we first start by considering an arbitrary Entomological Inoculation Rate ($EIR$, the number of infectious bites an individual receives each year) and Sporozoite Rate ($SR$, the prevalence of sporozoites in the mosquito population), which together define an overall annual biting rate ($ABR$).

$$ABR= \frac{EIR}{SR}$$

For the purposes of the results in the main text, we select an $EIR$ of 1 and an $SR$ of 0.05 to give an $ABR$ of 20, though we stress these choices are arbitrary and meant to be illustrative only, and that the methods below could be used to calculate the results for any combination of $EIR$ and $SR$. For a given $ABR$ and for each *An. stephensi* time-series $i$, this $ABR$ is proportionally divided up over the course of 365 days according to the normalised vector density at each timepoint, such that the biting rate $b$ for time-series $i$ on day $d$ is given by:

$$b_{i, d}= ABR\left( \frac{D_{i,d}}{\sum_{d=1}^{d=365} D_{i,d}} \right)$$

where $D_{i,d}$ is the normalised vector-density on day $d$ for time-series $i$. Because the sampling resolution of the studies we collated was never finer than monthly, we then use $b_{i, d}$ to calculate an average daily biting rate for each month, $b_{i, m}.$ $b_{i, m}.$ therefore describes the expected daily number of bites an individual receives in month $m$. The number of bites an individual would receive during a specific day of human landing catch sampling during month $m$ can then be considered a draw from a Poisson distribution with rate as follows:

$$C_{i,m} \sim Poisson\left( \lambda=b_{i, m} \right)$$

The expected number of mosquitoes caught over multiple days and months of mosquito sampling can then be calculated by exploiting the following property of the Poisson distribution:

$X_{1} \sim Poisson\left( \lambda_{1} \right)$ & $X_{2} \sim Poisson\left( \lambda_{2} \right)$

then $X_{1}+X_{2} \sim Poisson\left( \lambda_{1}+\lambda_{2} \right)$

Given this, for time-series $i$, carrying out mosquito sampling for $n_{m}$ consecutive months starting at month $j$, and within each month carrying out $n_{d}$ days-worth of sampling, the total number of *An. stephensi* expected to be caught is given as follows:

$$C_{i,j, n_{m},n_{d}} \sim Poisson\left( \sum_{m=j}^{m=(j+n_{m}-1)} n_{d}b_{m}) \right)$$

from which the probability of not sampling *An. stephensi* (i.e. the total number of *An. stephensi* caught is equal to 0) during that sampling period can be calculated.

For each time-series, we then identified the month in which monthly rainfall peaked, and the month in which vector density was highest (noting that these months were very rarely the same month). We then calculated the cumulative probability of *An. stephensi* detection under a range of different surveillance strategies. Specifically three strategies were simulated:

- **Vector-Peak Timed:** Starting the survey at the month with peak vector density (noting that in the absence of pre-existing detailed entomological information this is largely a hypothetical quantity, meant to illustrate the maximum detection probability that could be achieved).
- **Rainfall-Peak Timed:** Starting the survey at the month with peak rainfall.
- **Random Month Timed:** The expected cumulative probability of detection achieved if the survey was started during a random month (calculated in practice by simulating survey starting in each of the year’s 12 months and then calculating the average cumulative probability of these surveys).

In addition to varying the timing of the survey (which varies according to the surveillance strategy considered, as described directly above), we also varied the amount of sampling effort (number of days sampled within each month) and the overall duration of the (i.e. how many consecutive months were sampled). Note that the aim here is not to describe the exact probability of missing *An. stephensi* in any given entomological survey, as this will depend on a wide array of other, poorly defined and heterogeneous factors (such as type of catch methodology used etc). Instead, the aim is to highlight how variation in seasonal dynamics can influence the nature of surveillance required to successfully *An. stephensi*.

**Modelling of Malaria Transmission and the Impact of *Anopheles stephensi***

We integrated the temporal profiles of *An. stephensi* abundance into a well-established deterministic compartmental model of *Plasmodium falciparum* malaria transmission and disease^20–22^ to explore the implications of the vector’s establishment and seasonality on the dynamics of malaria transmission, with a particular focus on areas where malaria transmission is currently low or absent. What follows is a description of the mathematical modelling framework in general terms, followed by specific details about how exactly this framework was used to model malaria transmission underpinned by *An. stephensi* in settings where malaria is currently absent or only minimally present.

The deterministic malaria model used here considers both human and mosquito populations. Humans begin as Susceptible (*S*), and upon infection (at a rate which is dependent on the force of infection they experience), progress to either Asymptomatic (*A*) or clinical disease, with the comparative probability of these two outcomes depending on the degree of acquired natural immunity due to previous exposure to the parasite. If an individual progresses to clinical disease, they enter either a Treated (*T*) or Clinical Disease (*D*) state that depends on the probability of receiving treatment. For those treated, individuals progress through a period of prophylactic protection following treatment (*P*), and then return to the susceptible compartment. For those developing clinical disease, they remain symptomatic for the duration of the disease, before moving to an asymptomatic state (*A*, detectable by light microscopy), before subsequently moving to a submicroscopically infected state (U, not detectable by light microscopy). Individuals who are currently asymptomatically infected (including individuals in both the *A* and *U* states) can be reinfected and develop clinical disease once again – if this does not occur, they subsequently clear the infection and return to the susceptible state.

Adult mosquito populations and their preceding juvenile stages are also explicitly modelled. Immature mosquitoes start off as larvae, divided into early and late stage (*Es* and *Ls* respectively) which then mature into pupae (*P*) before eventually maturing into adult mosquitoes. Adult mosquitoes are further stratified according to infection with *P.falciparum* status – they begin as susceptible (*Sm*) and upon infection, progress to an exposed (but un-infectious, *Em*) state, and then onto the infectious state (*Im*) following the extrinsic incubation period (EIP, assumed to be constant over time). Mosquitoes are infected through exposure to humans currently possessing transmissible infections i.e. the treated (*T*), clinical disease (*D*), asymptomatic (*A*) and submicroscopic (*U*) infection states.

Seasonality in mosquito abundance is incorporated through a flexible, time-varying carrying capacity that in broad terms describes temporal variation in the ability of the local environment to support mosquito breeding. The value of this carrying capacity relative to the size of the mosquito population influences the mortality of early and late-stage larvae, which as previous modelling work has shown, enables the model to accurately and adequately capture temporal fluctuations in mosquito abundace^23^. We integrate each of the seasonal profiles of *An. stephensi* density implied by the corresponding time-series of catch data into the model, matching the carrying capacity to the empirically observed temporal variation in *An. stephensi* abundance. Estimates of the bionomic properties of *An. stephensi* (specifically the mosquito’s daily mortality, degree of anthropophagy, degree of endophily and the proportion of bites taken on individuals indoors and/or in bed) were taken from previous work that reviewed the properties^24^, and the vector to human ratio arbitrarily set to 20, which corresponds to approximately 9% malaria prevalence in a setting where the risk of malaria is constant year round (i.e. a perennial setting). Indoor residual spraying (IRS) is assumed to reduce malaria burden primarily by both killing adult mosquitoes and deterring them from biting and feeding – specifically, IRS can either repel before biting and feeding, or kill following biting (when the vector rests on a sprayed wall). The efficacy of IRS decays over time due to a loss of insecticide. The efficacy of the different IRS compounds considered (bendiocarb, clothiandin and pirimiphos methyl), as well as the different rates of efficacy decay were parameterised using Sherrard-Smith et al 2018^25^. We modelled the impact of a single round of IRS, timed according to a range of different strategies that largely mirror the strategies described in the section on surveillance and entomological monitoring above. Specifically, these were:

- **Optimal-Timing:** Starting the survey at the timepoint where the reduction in incidence is maximised (noting that in the absence of pre-existing detailed entomological information on the timing of peak vector abundance, this is a hypothetical quantity, meant to illustrate the maximum impact that could be achieved with perfect information).
- **Rainfall-Peak Based Timing:** Starting the survey at the midpoint of the month with peak rainfall.
- **Random Month:** The expected reduction in malaria incidence achieved if the IRS campaign was started during a random month (calculated in practice by simulating survey starting in each of the year’s 12 months and then calculating the average cumulative probability of these surveys).

In all cases, the impact was calculated by comparing the reduction in malaria burden (as measured by total annual incidence in the 12-month period following spraying) compared to a counterfactual of no IRS.

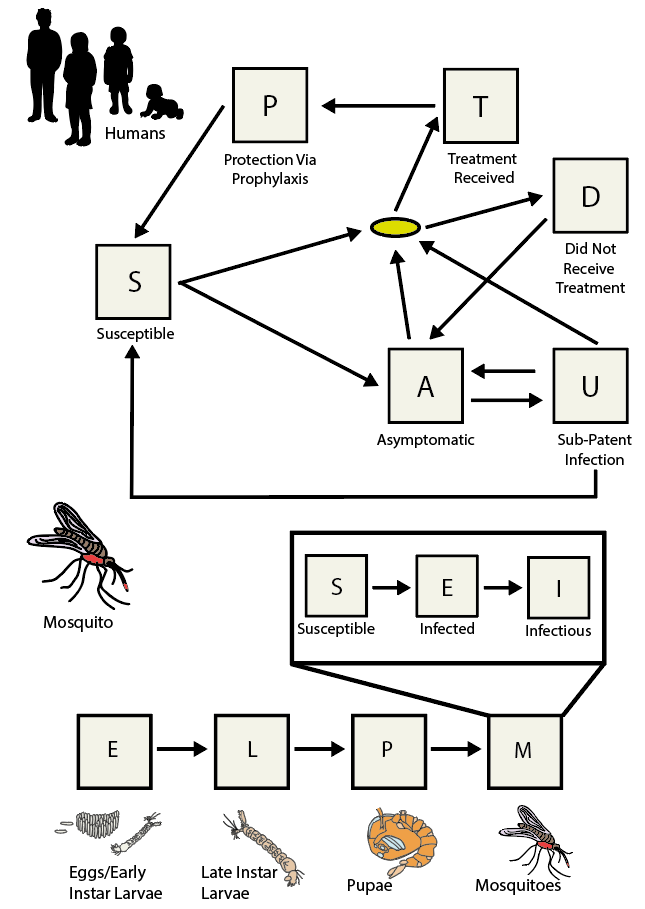

**Supplementary Figure Model Schematic:** Humans exist in either *S* (Susceptible), *A* (asymptomatic infection), *T* (infected and treated), *D* (infected and have clinical disease), *U* (submicroscopically infected) or *P* (prophylactically protected from infection by treatment received). The full-life cycle of the mosquito is modelled, with states including *E* (eggs/early larvae), *L* (late instar larvae), *P* (pupae), and finally *M* (mature adult mosquitoes). Mosquitoes in the *M* state begin in state *S* (susceptible) and upon infection more to a latently infected (but not yet infectious state) denoted by *E*. Upon becoming infectious they transition to the *I* state. Arrows show transitions between states, with the yellow oval indicating a decision point in the human part of the model based on whether the individual receives treatment or not.

**Supplementary Information 3: Additional Figures and Results**

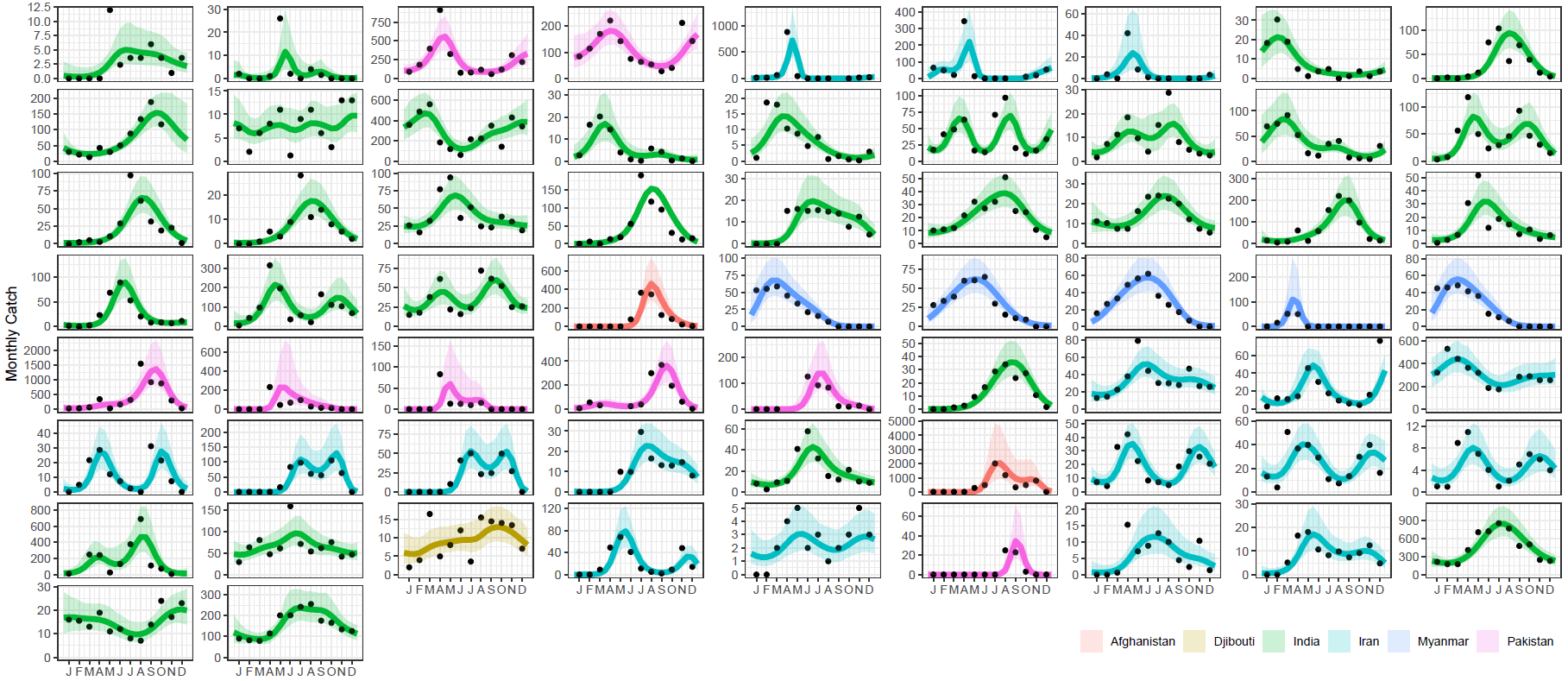
**Supplementary Figure 1: Results of model fitting to the longitudinal entomological data collated in this study.** Reviews of the literature in tandem with previously published databases of entomological data identified 65 *Anopheles stephensi* time-series matching the inclusion criteria (>10 months of catch data at monthly temporal resolution or finer), and a negative binomial gaussian process with period kernel fitted to each time-series. For the results presented above, black points are the data, and the lines represent the model output, coloured according to the country in which the study was conducted. Line indicates the mean model output, with the shaded ribbon delineating the 95% credible interval (CI).

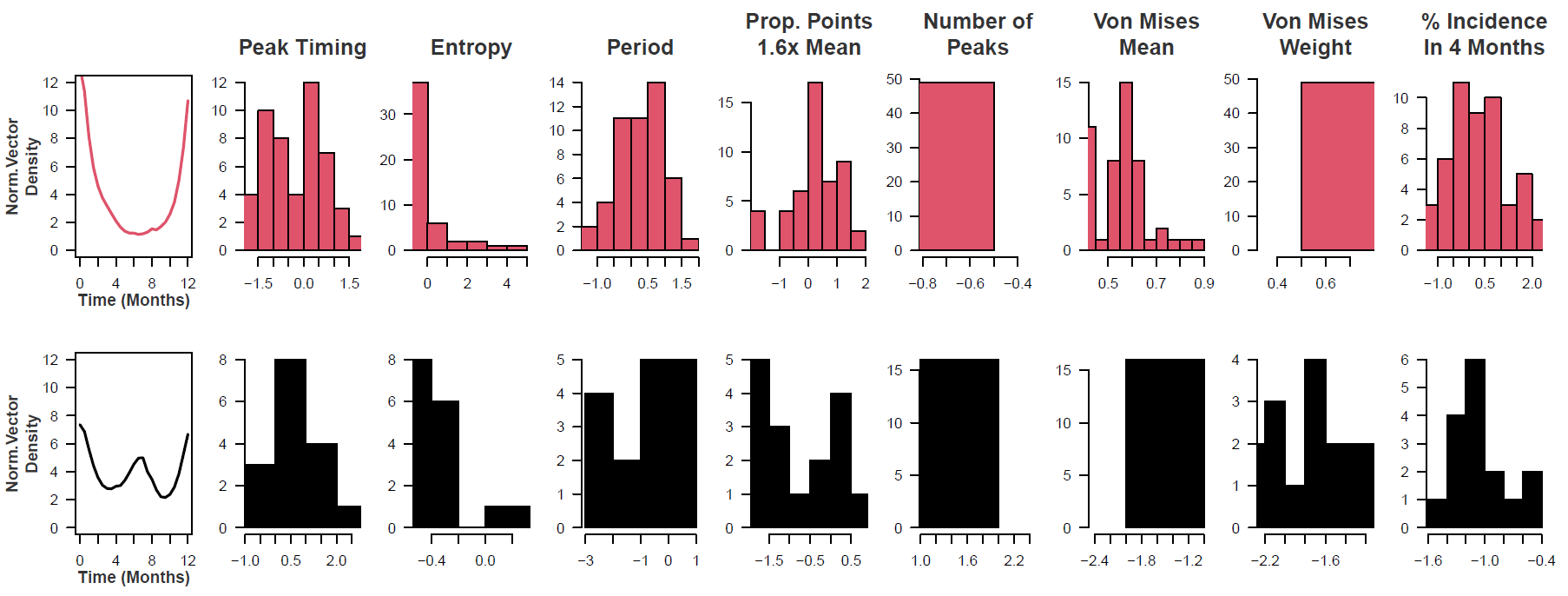

**Supplementary Figure 2: Archetype/Cluster Temporal Properties.** A series of mathematical operations were applied to the fitted time-series to characterise and explore their temporal properties. The results of this characterisation were then clustered using the k-means algorithm. For each cluster, the mean temporal profile is displayed, as well as the underlying distribution of values for each temporal property for each cluster (where the values for a given temporal properties for all time-series have first been normalised and standardised to have mean 0 and unit variance). For further information on each of these operations, see ***Supplementary Information: Time Series Characterisation and Analysis.***

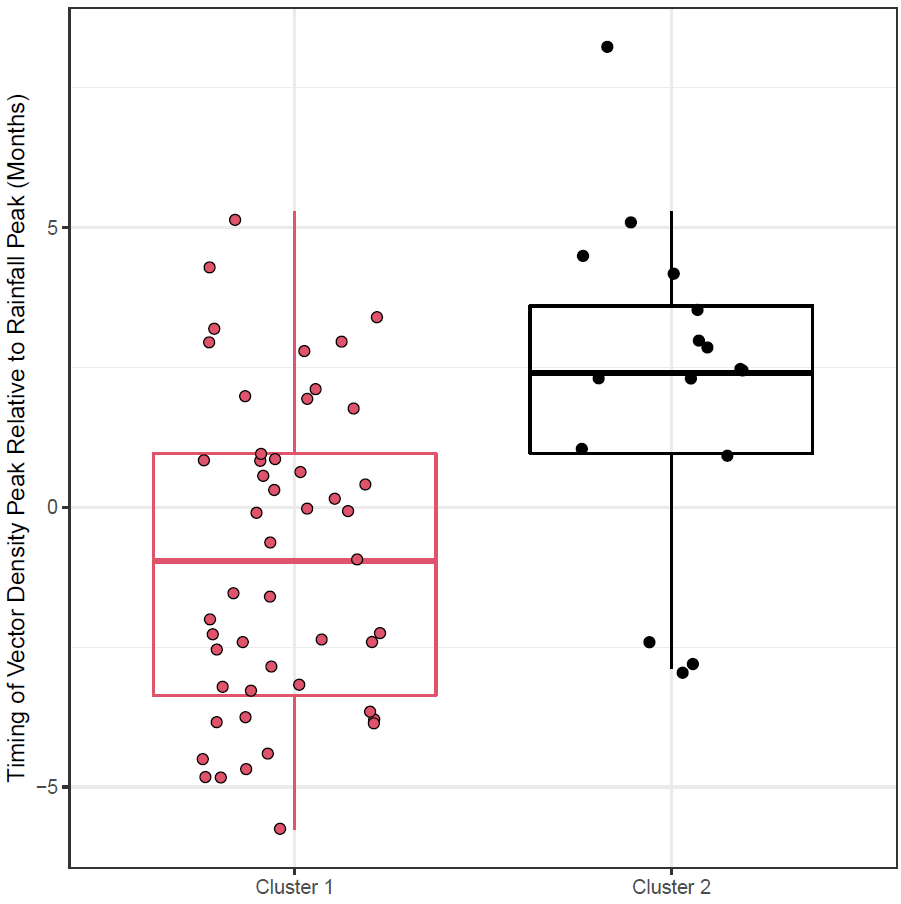

**Supplementary Figure 3: Timing of Vector Density Peak Relative to Rainfall Peak, Stratified by Cluster.** For each study location and time-series, we calculated the time-difference (in months) between the peak vector density and peak monthly rainfall. Our results highlighted systematic differences between clusters, but also significant variation within clusters.

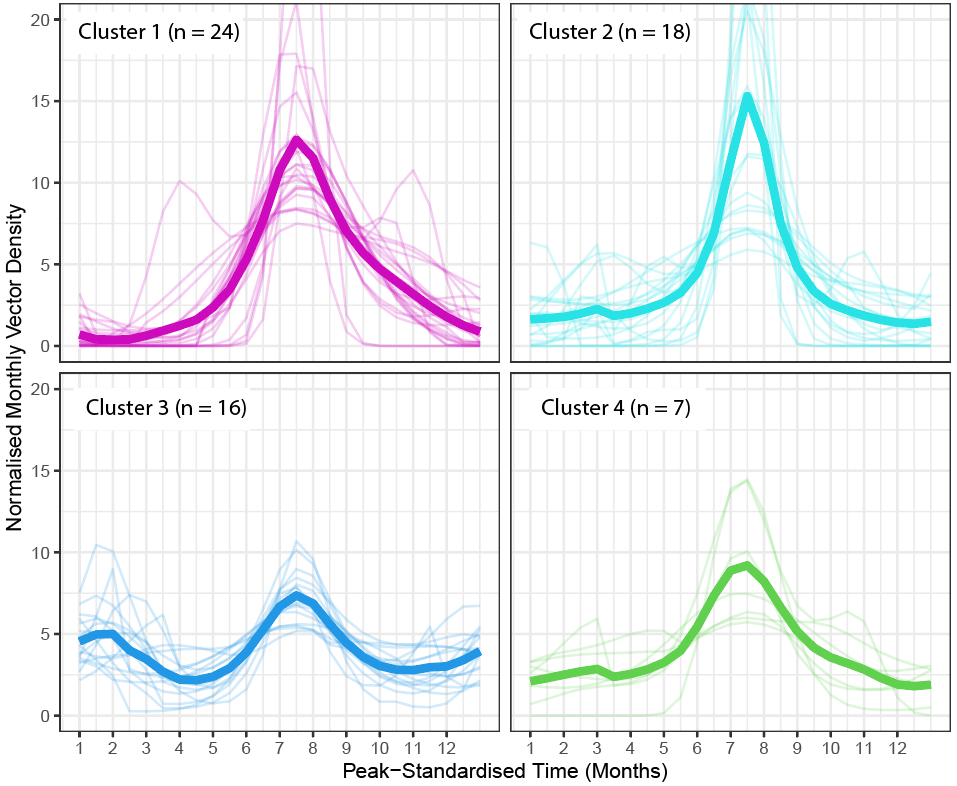

**Supplementary Figure 4: Results of Clustering For 4 Clusters Instead of 2.** In order to further investigate the different patterns of temporal dynamics present in the collated dataset, we re-ran the k-means clustering algorithm this time specifying 4 clusters. The less seasonal cluster from the 2 cluster analysis in the main text (Cluster 2 in the main text results) was retained (here Cluster 3), and Cluster 1 from the main text was further disaggregated into 3 different clusters (here, Clusters 1, 2 and 4), each defined by different peak timings (mean timing of vector density peak 7, 8.25 and 5.86 months after January for Clusters 1, 2 and 4 respectively) and the timing of the vector peak relative to peaks in rainfall (rainfall peak on average 1.03 and 2.32 months before vector density peak for Clusters 1 and 2, 1.09 months after vector density peak on average for Cluster 4).

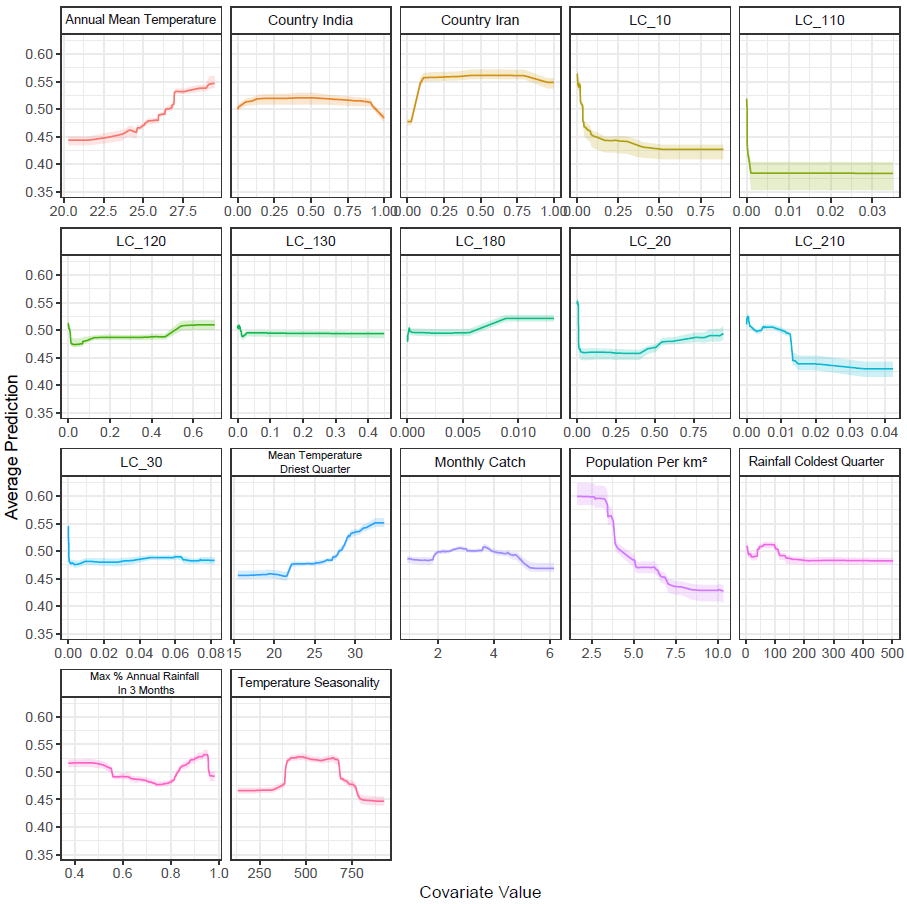

**Supplementary Figure 5: Partial Dependence Plots for Covariates Used in the Random Forest Classification Modelling.** The y-axis on the left shows the probability of the time-series belonging to Cluster 2 (i.e. a high probability indicates the time-series is predicted to likely belong to Cluster 2, a low probability indicates the time-series likely belongs to Cluster 1). The x-axis describes the value of the (scaled, normalised) covariate.

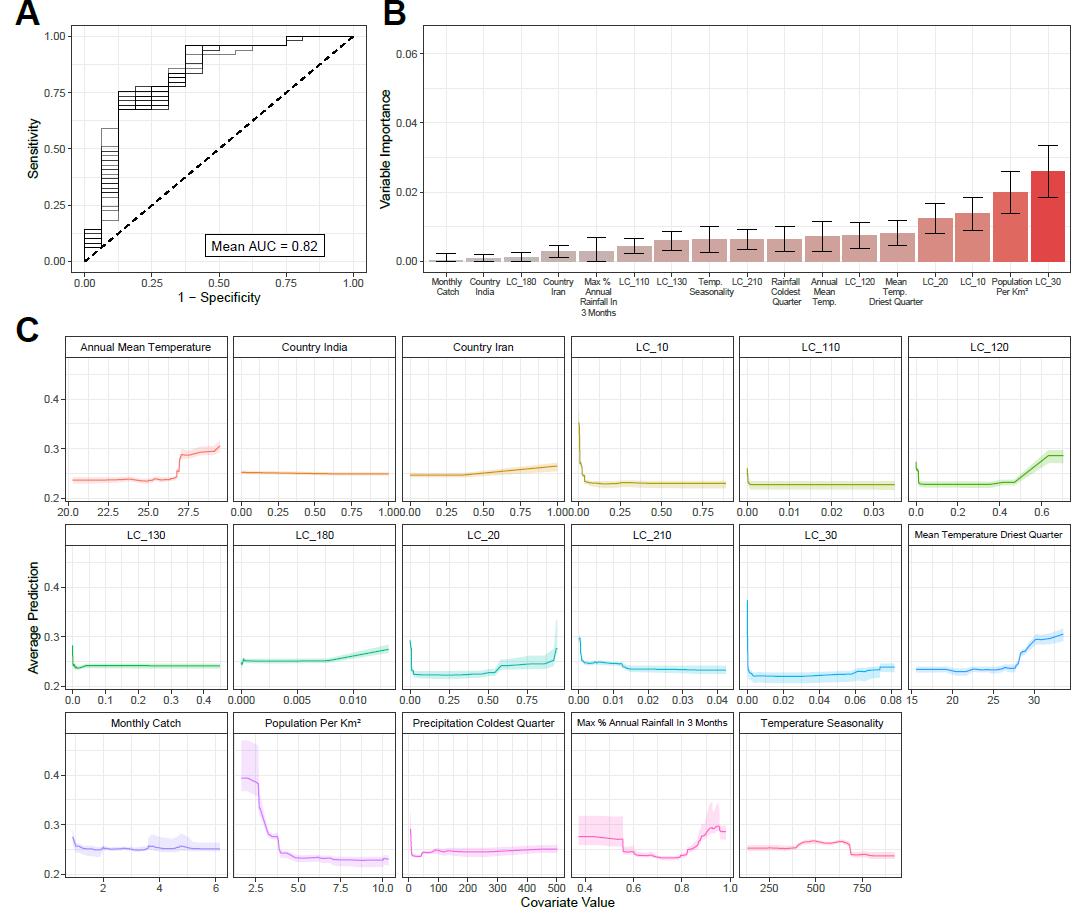

**Supplementary Figure 6: Random Forest Classification Results Without Upsampling Cluster 2.** Due to the extreme class-imbalance of Clusters 1 and 2 (49 vs 16 time-series respectively), the results presented in the main text are following upsampling of the Cluster 2 time-series to create a dataset with equal numbers of time-series belonging to each cluster. As a sensitivity analysis, we also carried out the random forest fitting without upsampling and assessed both model fit (as measured by AUC, **(A)**) and variable importance **(B)**. Model performance was somewhat reduced compared to the upsampled data (mean AUC of 0.81 vs mean AUC >0.9 for the upsampled dataset), whilst variable importance results were broadly consistent across both analyses, with population per square kilometre and various land-cover measures all emerging as important predictive variables. We also present partial dependence plots for all of the included covariates **(C)**. The y-axis on the left shows the probability of the time-series belonging to Cluster 2 (i.e. a high probability indicates the time-series is predicted to likely belong to Cluster 2, a low probability indicates the time-series likely belongs to Cluster 1). The x-axis describes the value of the (scaled, normalised) covariate.

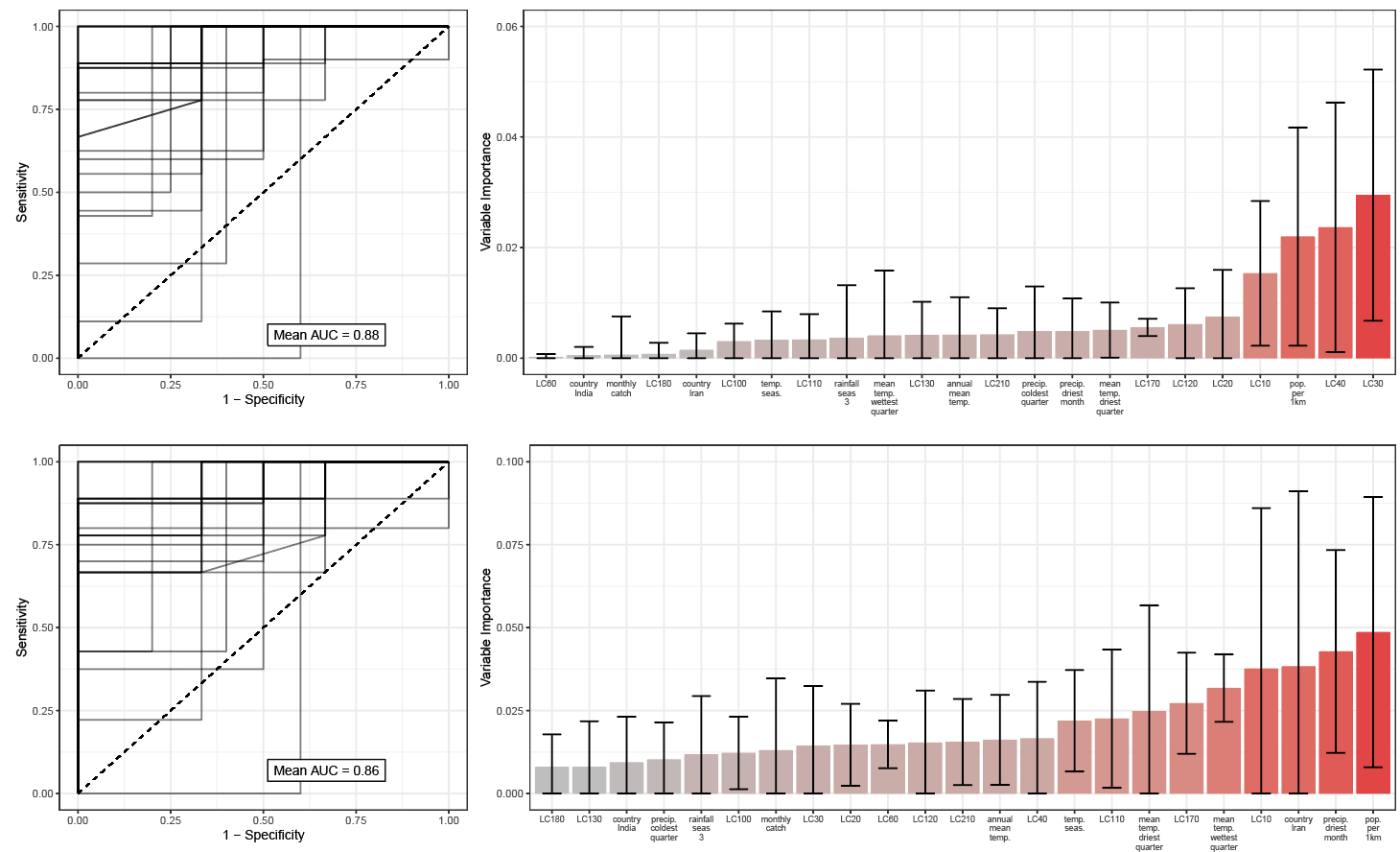

**Supplementary Figure 7: Random Forest Classification Results With Hold-Out Data.** Due to the overall sample size (n = 65 time-series), the results presented in the main text were generated using a random forest-based workflow where final model fitting (using hyperparameters tuned using 6-fold cross-validation) utilised the entirety of the dataset. As a sensitivity analysis, we also carried out the random forest fitting holding out a small portion of the dataset (n = 9) during model fitting, with model performance subsequently evaluated on this held-out data. Results presented above are in the case where data was upsampled to address class imbalance (top) and where no upsampling was carried out (bottom).

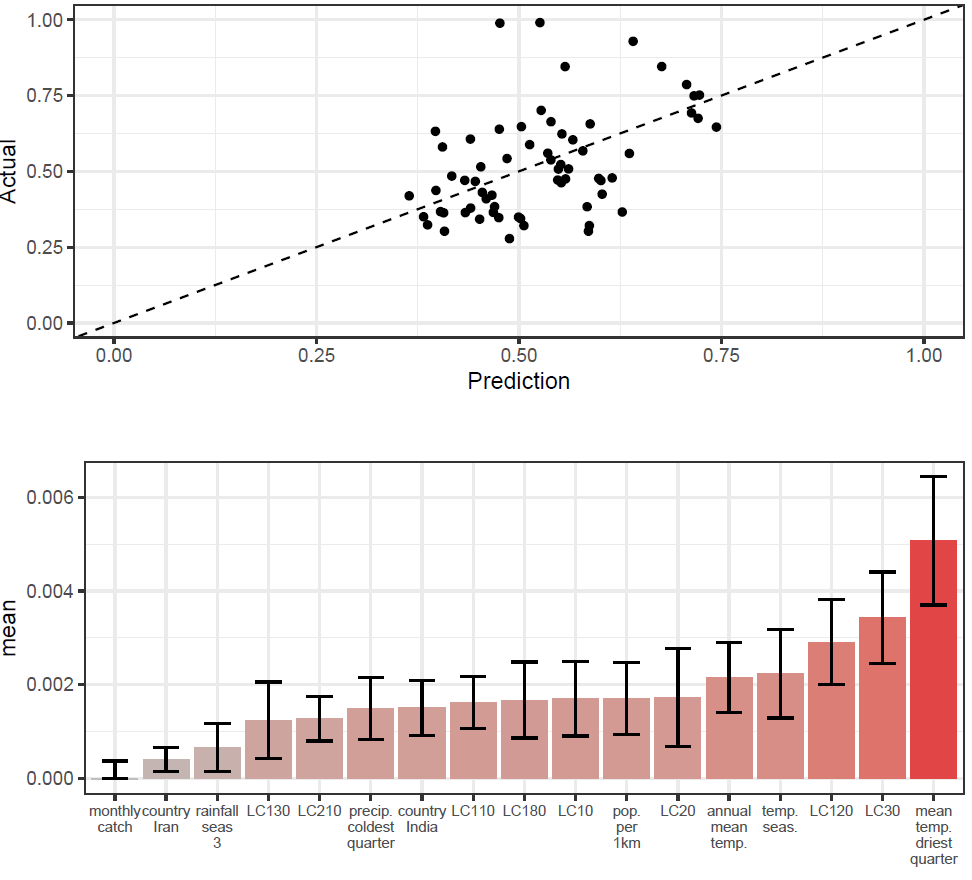

**Supplementary Figure 8: Random Forest Prediction of Percentage of Vector Density In Any 3 Month Period.** As a further sensitivity analysis, we used a random forest modelling approach to predict the percentage of vector density occurring in a single continuous 3-month period. Results presented above are the average of 25 independents random forest model fittings, with no upsampling of the data carried out, and the final model fitted (using hyperparameters tuned using 6-fold cross-validation) to the full dataset. Model predictive power was moderate, with correlation between predicted and actual values = 0.43.

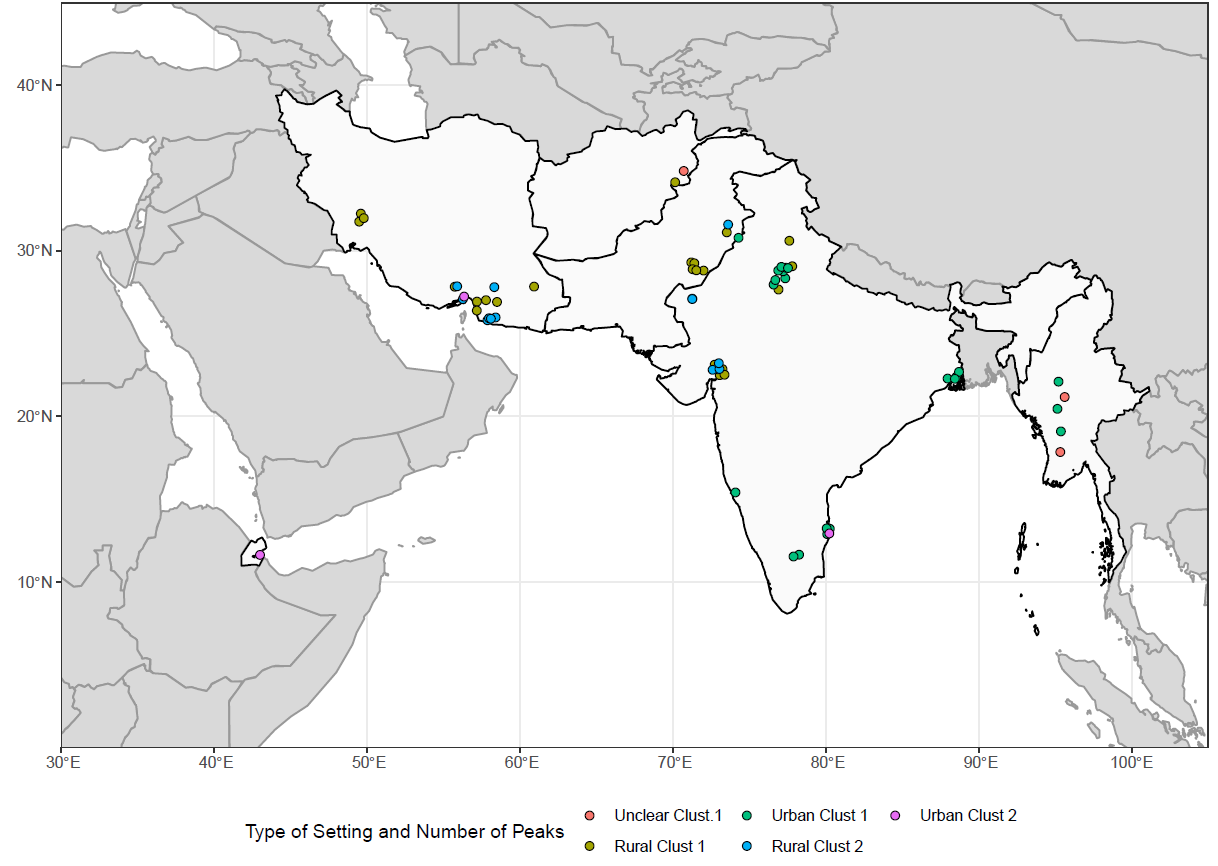

**Supplementary Figure 9: Sources and Locations of *Anopheles stephensi* Time-Series Data According to Urban/Rural Assignment.** Collated time-series are displayed above coloured according to 1) whether or not the study was carried out in an urban or rural location; and 2) which cluster they were assigned to.
